# Glucocorticoid-driven gene expression in circulating monocytes and neutrophils in health and severe inflammation

**DOI:** 10.1101/2023.05.10.23289779

**Authors:** Arthur Molendijk, Leo Koenderman

## Abstract

Glucocorticoids (GCs) are used as anti-inflammatory and immunosuppressive drugs in many immune mediated diseases, but their use in sepsis and shock is controversial. This is caused in part by a lack of information regarding the responding cell types and GC-regulated genes *in vivo*. We used public blood transcriptomic datasets and GC-induced query genes to obtain 2 robust gene expression correlation signatures of GC induction, either in the absence or in the presence of severe inflammation. GC signature 1 originated from circadian cortisol with biases for gene expression in NK cells and neutrophils. GC signature 2 originated from GC in severe inflammation, mainly with biases for gene expression in monocytes and neutrophils. Many genes upregulated by GC treatment in septic shock and burn shock were also present as high-ranking genes in GC signatures, which pointed to their direct regulation by GC. Robust GC signatures were also obtained from dataset collections of monocytes and neutrophils, separately, and predicted cellular effects. Additionally, gene induction by GC was put into a wider framework of gene expression in circulating monocytes and neutrophils in health and systemic inflammation. We present and interpret a large number of GC-regulated genes in different blood cells and tissues, and select 2 whole blood transcriptomic biomarker gene sets, GC-1 and GC-2, for monitoring cortisol action in health, and in severe inflammation, respectively. GC signature 2 was found in sepsis and many other inflammatory diseases, both from treatment with GC, and from endogenous GC.

## INTRODUCTION

### Use of glucocorticoids in therapy

Endogenous cortisol links the endocrine and immune system, and has an important role in regulating inflammatory events. Synthetic glucocorticoids (GCs) are widely used in medicine because of their strong anti-inflammatory and immunosuppressive mode of action. Systemic corticosteroid therapy is routinely used in autoimmune diseases such as rheumatoid arthritis (RA) and systemic lupus erythematosus (SLE), during flares in MS, during severe exacerbation in pulmonary diseases such as COPD and asthma, and as cortisol replacement in Addison’s disease (Chan et al., 2020; Johannsson et al., 2015; Reichardt et al., 2021; Strehl et al., 2019). GCs with reduced systemic bioavailability are also used in active inflammatory bowel disease (IBD) (Dubois-Camacho et al., 2017). A controversy is present concerning the use of GCs in the treatment of acute inflammation such as found during septic and burn shock as well as in ARDS and COVID-19 (Djillali Annane et al., 2017; Briegel et al., 2018; Carlet et al., 2020; Chan et al., 2020; Vandewalle & Libert, 2020; Venet et al., 2015). Clinical trials have shown a beneficial effect of GC treatment in shortening septic shock duration, but inconsistent results have been published regarding survival at 180 days (Annane et al., 2018; Venkatesh et al., 2018). The use of GC as additional therapy in septic shock is now recommended if vasopressor therapy is not effective enough in maintaining target blood pressure (Evans et al., 2021). Considering the physiological importance of endogenous GCs and the wide use of systemic GC therapy, biomarkers for monitoring GC activity at the cellular level are very useful (Alder et al., 2018; Chan et al., 2020; Russell, 2018). Blood transcriptomics is an important technology for obtaining such biomarkers, and has been used to develop an 8 gene biomarker of GC exposure especially in RA and SLE (Hu et al., 2018). Blood transcriptomics has also been used to classify heterogeneity in sepsis (Leligdowicz & Matthay, 2019; Tsakiroglou et al., 2023), but without using GC-induced genes, as biomarkers, to assist in classification. Furthermore, the use of GC-induced genes for monitoring the cellular effects of GC therapy in septic and burn shock has not been adequately applied.

### Objective

Our objective here was to use large collections of blood transcriptomics datasets to obtain GC-driven gene signatures in the absence or presence of severe systemic inflammation, such as found in sepsis, ARDS, and trauma. Secondly, profiling signature gene expression based on cell type and inflammatory state was performed to help select suitable biomarkers for monitoring cortisol activity in different illnesses.

### Cortisol, GR, and cellular effects

Cortisol is a steroid hormone and the major end product of the HPA (Hypothalamic Pituitary Adrenal) axis, which plays an important role in the body’s stress response. Different stresses, both physical or emotional, stimulate the HPA axis and thereby transiently increase systemic ACTH and cortisol concentrations (Herman et al., 2016; Oster et al., 2017). Moreover, circadian stimulation of the HPA axis results in increasing cortisol concentration during late sleep, before sunrise, peaking around the sleep wake transition at 6h-10h, and bottoming in the evening, early night between 22h-2h (Oster et al., 2017). Changes in cortisol concentration affect blood cell trafficking and mobilization, resulting in stress and circadian rhythm related shifts in different immune cell percentages (Ince et al., 2019; Olnes et al., 2016; Sautron et al., 2015; Shimba & Ikuta, 2020). In sepsis and critical illness more generally, the central HPA axis is first transiently activated, but then becomes suppressed as indicated by low plasma ACTH concentration. However, plasma cortisol levels remain elevated partially due to increased production and mainly due to decreased breakdown in liver and kidney (Boonen et al., 2013; Langouche et al., 2023; Van den Berghe et al., 2022). Cortisol availability is further increased by different peripheral adaptations such as decreased hepatic synhesis of GC carrier proteins resulting in more free cortisol. Intracellular cortisol availability in target tissues might also be increased by enhanced inactive cortisone to cortisol enzymatic conversion (Langouche et al., 2023; Van den Berghe et al., 2022). Early research indicated that high cortisol levels occurring in septic shock might still be inadequate in relation to the severity of the disease (Annane et al., 2000). This insufficiency was referred to as CIRCI (critical illness-related corticosteroid insufficiency), and was caused by adrenal insufficiency together with tissue corticosteroid resistance, and characterized by a strong proinflammatory response (Marik et al., 2008). Presently, CIRCI is hard to diagnose, but might still require treatment with GC (D. Annane et al., 2017; Djillali Annane et al., 2017). Recently, it has been argued that a form of CIRCI might only occur in prolonged critical illness (Téblick et al., 2022), and that CIRCI is a poor rationale for the use of GC in septic shock (Langouche et al., 2023; Téblick et al., 2022; Van den Berghe et al., 2022). In this view, high doses of GC in septic shock would not remedy insufficiency due to GC resistance in CIRCI, but improve blood pressure as a pharmalogical effect (Langouche et al., 2023; Van den Berghe et al., 2022).

Natural (cortisol in humans) and synthetic (e.g. dexamethasone) GCs are lipophilic molecules, that can traverse cell membranes. Cellular effects of GCs can be genomic (transcriptional) mediated by the glucocorticoid receptor (GR), or non-genomic (transcription independent), involving GR or other putative GC receptors (Escoter-Torres et al., 2019; Timmermans et al., 2019). In the absence of GC ligand, the glucocorticoid receptor (GR) is a part of a matured protein heterocomplex in the cytoplasm (Escoter-Torres et al., 2019; Petta et al., 2016; Timmermans et al., 2019). Upon ligand binding, GR translocates to the nucleus, and binds as a monomer, dimer or tetramer to half site or pseudo-palindromic GREs (glucocorticoid response elements) in DNA, which are situated in promoters and enhancers, to activate its target genes (Escoter-Torres et al., 2019; Gerber et al., 2021; Johnson et al., 2021; Petta et al., 2016; Strickland et al., 2022; Timmermans et al., 2019; Vettorazzi et al., 2021). Important GR gene targets for activation produce anti-inflammatory factors, such as DUSP1/MKP-1, NFKBIA/IkBalpha, TSC22D3/GILZ, and TNFAIP3/A20 (Oh et al., 2017). GR physically interacts with c-Jun of AP-1 (Diamond et al., 1990; Yang-Yen et al., 1990), and with the p65 subunit (RelA) of NF-kB (Caldenhoven et al., 1995; Ray & Prefontaine, 1994; Scheinman et al., 1995). These protein-protein interactions are important for mutual antagonism between anti-inflammatory GR and inflammatory NF-kB and AP-1 activity, as reviewed (Petta et al., 2016). GR also functions in repression of gene transcription, which requires direct DNA binding of the GR to GREs (Escoter-Torres et al., 2020). The exact mechanisms of gene repression, including a possible role of tethering of GR to DNA-bound AP-1 or NF-kB, are being investigated (Escoter-Torres et al., 2019; Fadel et al., 2023; Gerber et al., 2021; Strickland et al., 2022; Vettorazzi et al., 2021). Many effects of GCs on neutrophils, monocytes, and monocyte-derived macrophages, are thought to relate to the cell biology of different phases of the local inflammatory response to injury and infection, where a pro- inflammatory phase triggered by DAMPS and PAMPS, is followed by a resolution phase, and a tissue repair phase (Desgeorges et al., 2019; Ehrchen et al., 2019; Ronchetti et al., 2018).

GCs affect monocytes, and monocyte-derived macrophages depending on their differentiation and activation status, suppressing the pro-inflammatory phenotype, and stimulating the anti- inflammatory phenotype by increasing their phagocytosis of apoptotic cells (Desgeorges et al., 2019; Ehrchen et al., 2019). Phenotypic effects of GCs on immune cells can be deduced, to some degree, from their transcriptomes and epigenomes. Transcriptomes of different isolated blood cells and primary cell types, treated *in vitro* with GC, are highly cell type- dependent, and predict testable phenotypic effects (Franco et al., 2019). Such transcriptomes are potentially valuable data for development of selective therapies (Buttgereit, 2021; Franco et al., 2019), which would also be true for cell specific transcriptomes induced by GC *in vivo*, as accessible by a gene expression correlation approach.

### Trauma and sepsis

Protective responses to mild or moderate trauma are thought to become exaggerated and destructive after massive trauma, of the type that was unsurvivable before the advent of medicine (Lord et al., 2014). Severe trauma leads to rapid systemic inflammation, which is excessive and commonly referred to as SIRS (systemic inflammatory response syndrome). Pro- and anti-inflammatory processes occur together, as studied at the level of blood plasma cytokines/chemokines and the leukocyte transcriptome (Xiao et al., 2011). Patients who survive SIRS and early MOF (multiple organ failure) are still at low risk of developing PICS (persistent inflammation, immunosuppression and catabolism syndrome) (Gentile et al., 2012; Hesselink et al., 2020). After severe injury, systemic exposure to DAMPS leads to activation of complement and coagulation pathways, and to activation of innate immune cells.

Therefore, trauma can lead to remote organ injury and sepsis (Huber-Lang et al., 2018; Lord et al., 2014; Pottecher et al., 2019). Sepsis is defined as a life-threatening organ dysfunction caused by a dysregulated host response to infection (Singer et al., 2016). The degree of organ dysfunction is represented by a SOFA (sequential organ failure assessment) score (Singer et al., 2016). Patients who survive early multiple organ failure from SIRS in sepsis often develop chronic critical illness involving PICS (Darden et al., 2021). For therapy of sepsis, the immunology of the host response is very important besides the nature of the infection (Jarczak et al., 2021). Sepsis is heterogenous, for example in the relative presence of hyperinflammation and immune suppression which may also change over time, and a major effort is ongoing in patient stratification, including the use of blood leucocyte transcriptomics, to possibly improve effects of standard and future therapies (Leite et al., 2024; Leligdowicz & Matthay, 2019; Pelaia et al., 2023; Stanski & Wong, 2020; Tsakiroglou et al., 2023; van der Poll et al., 2021; Van Der Poll et al., 2017). Sepsis starts with detection of PAMPs from pathogens, and DAMPs from tissue damage, causing a systemic inflammatory response.

Several pro-inflammatory mechanisms become deregulated including activation of complement and coagulation, immune cells, endothelial cells, and platelets. On the other hand, immunosuppressive responses are induced in parallel including lymphocyte apoptosis, reduced antigen presentation by APCs, and macrophages shifting to an alternative (M2) activation state (Hotchkiss et al., 2016; Jarczak et al., 2021; van der Poll et al., 2021; Van Der Poll et al., 2017; Wiersinga & van der Poll, 2022).

### Monocytes and neutrophils in severe inflammation

Low surface expression of HLA-DR on blood monocytes is an important marker for immunosuppression in sepsis, in surgery, and in cancer (Hibbert et al., 2018; Venet & Monneret, 2018), and correlates negatively with serum cortisol concentrations (Kim et al., 2010; Mengos et al., 2019; Tulzo et al., 2004). Monocytes found in sepsis are more phagocytic, and less responsive to endotoxin stimulation (endotoxin tolerant), but express higher levels of cytokines and interleukins compared to monocytes from healthy controls (Shalova et al., 2015). They display an activation pattern that is similar as found in macrophages as judged by gene expression analysis (Liepelt et al., 2020; Washburn et al., 2019). Recently, single cell sequencing has allowed a better description of monocyte heterogeneity in sepsis, ARDS, and COVID-19, and different monocyte cellular states have been identified, some of which are strongly expanded (Jiang et al., 2020; N. Liu et al., 2021; Reyes et al., 2020; Reyes et al., 2021; Schulte-Schrepping et al., 2020; Wang et al., 2021; Wen et al., 2020). In case of COVID-19, a gene expression signature of GC treatment has been identified for monocytes (Knoll et al., 2024), but not yet for ‘sepsis monocytes’.

Neutrophils in severe systemic inflammation, such as found after trauma and in sepsis, are dysfunctional which is associated with the susceptibility to infections, and cause of tissue damage. The functions of neutrophils in acute inflammation act as a double edged sword. At the one hand, neutrophils are essential in pathogen defense and tissue repair, but on the other hand deregulated neutrophils are involved in collateral damage to host healthy tissue (Hesselink et al., 2019; Kovtun et al., 2018; Leliefeld et al., 2016; Mortaz et al., 2018).

Neutrophils isolated from the blood of sepsis or ARDS patients are functionally altered including reduced chemotaxis, and delayed apoptosis. Interestingly enough no change in phagocytic capacity was found for these cells (Demaret et al., 2015; Juss et al., 2016).

Different neutrophil populations occur in severe systemic inflammation, based on cell surface markers, nuclear shape, cellular density, and functional characteristics (Hellebrekers et al., 2018; Hesselink et al., 2019; Leliefeld et al., 2016; Mortaz et al., 2018; Pillay et al., 2012; Shen et al., 2017). Severe systemic inflammation results in the appearance of low density neutrophils (LDNs), with different origins, including degranulated neutrophils, immature neutrophils, granulocytic MDSCs, and pro-inflammatory/primed LDNs (Hesselink et al., 2019). Transcriptomic studies, on bulk neutrophils or single cells in systemic severe inflammation including severe COVID-19 (Aschenbrenner et al., 2021; de Kleijn et al., 2012; Demaret et al., 2015; Juss et al., 2016; Kwok et al., 2023; Kwok et al., 2020; Reusch et al., 2021; Schulte-Schrepping et al., 2020; Sinha et al., 2022; Vieira Da Silva Pellegrina et al., 2015) have not evaluated the identification of co-expressed GC-induced genes.

### Result, importance

Here we used a meta-analysis to overcome some of the limitations of identifying glucocorticoid-induced genes in single blood transcriptomics datasets. Different GC inducible query genes were used on defined dataset collections, applying an expression correlation search to obtain large, robust signatures of glucocorticoid-driven gene expression. Signature genes were profiled for cell type-dependent expression, induction by GC *in vitro*, as well as for circadian expression, and upregulation during mild and severe inflammation. Signature 1 of glucocorticoid-driven gene expression in blood in the absence of severe inflammation, was largely derived from responses to circadian cortisol in NK cells and neutrophils. A set of genes (GC-1) selected from this signature was suitable for use as a blood transcriptional biomarker for the involvement of cortisol in the absence of severe inflammation. Signature 2 of glucocorticoid-driven gene expression in blood in severe inflammation, mainly originated from “severe inflammatory” neutrophils, among others characterized by upregulation of ARG1 encoding arginase 1, and from monocytes including M2 alternatively activated macrophage-like monocytes. Many GC signature genes were specifically upregulated upon GC treatment in septic and burn shock. A selection of genes (set GC-2) was used to demonstrate the presence of GC signature 2 in a wide range of inflammatory illnesses, originating both from exogenous and endogenous GC, which indicated a general importance of GCs in immunosuppression. This analysis extends the description of GC action on different immune cells and tissues *in vivo*, and gene set GC-2, derived from GC signature 2, will potentially be a useful biomarker for stratification of sepsis and other inflammatory diseases, and for monitoring of cellular responses to GC in severe inflammation.

## METHODS

### Datasets

This study made use of data in the public domain. Microarray gene expression data were downloaded from the NCBI GEO database (Barrett et al., 2013) and loaded into R, using Biobase and GEOquery packages from the Bioconductor software repository (Huber et al., 2015; Sean & Meltzer, 2007). Expression matrices from GSE expression sets were put into GEO DataSet (GDS) format-like expression tables (i.e. dataframes with ID_REF and IDENTIFIER columns for platform probe ID and gene symbol, respectively, and followed by sample value columns). If gene symbols were not directly provided with the platform, other platform probe identifiers were used to obtain matching gene symbols, via HGCN custom downloads, using the approved gene symbols from HGNC (Seal et al., 2023). RNA- sequencing phenotypic data were downloaded from NCBI GEO using GEOquery. The corresponding expression data were downloaded from the GEO website, and put into a GDS expression table-like format. Processed microarray and RNA-seq expression data were also downloaded from the ArrayExpress website (Athar et al., 2019), and read into R, along with the corresponding sample and data relationship (sdr) files, and array design files. Microarray and RNA sequencing data from ArrayExpress were also put into a GDS expression table-like format, using HGCN custom downloads in case of missing gene symbol columns in expression tables or platform.

### Dataset collections

Collections of datasets were assembled based on experimental description as provided in the accompanying metadata. Dataset collections varied in size, from whole blood in the absence of severe inflammation (40 and a subset thereof, 15), whole blood in the presence of severe inflammation (38 and a subset thereof, 15) (Table S1, sheet 15) to smaller sizes in case of separated monocytes and neutrophils (6-11), PBMCs (7), and tissues muscle, skin, and fat (5- 11). (Table S3, sheet 13). Gene expression tables, or subsets thereof, were left unchanged, without transformation of the original values except in a few cases where mentioned in results and supporting information for different collections. The original values mostly came in either base 2 logarithmic or linear scales. Single microarray platform IDs correspond with probe in Illumina or probeset in Affymetrix datasets, and are hereafter named probe for simplicity. The number of probes present in the ID-REF column corresponding to a single gene varies depending on microarray platform, whereas in case of the RNA-seq expression tables, each gene was uniquely represented by a single observation (row).

### Single gene expression correlation profiles from single datasets in a collection

Pearson correlation coefficients between the different probe expression values corresponding with a single query gene and all other gene probes in an expression table were calculated. A given query gene was used to obtain ranked expression correlation profiles for different datasets in a collection using the corresponding gene query probes present in each gene expression table from a collection. The number of profiles acquired per query gene depended on the presence of genes and corresponding probe numbers on dataset platforms.

### Averaging correlation coefficients in meta-analysis

Pooling correlation coefficients in meta-analysis, as used here of Pearson correlation coefficients of expression values for a pair of genes in different transcriptomic datasets, can be performed differently; either after Z transform, weighting, taking mean, and transformation of weighted mean value back to average correlation coefficient according to Fisher, or by taking mean of weighted correlations coefficients directly (Alexander, 1990; Field, 2005).

Weighing of different studies is applied according to within and between study variances, or according to study sample size, depending on method. Both methods are sufficiently accurate in obtaining an estimate, when population correlations vary (Field, 2005). Here, average correlation coefficients were directly taken as mean, without weighing by sample size, not to give too much weight to a few large datasets, and to have equal contributions of all datasets, which are potentially unique with respect to biased sampling of illnesses and other conditions. Average correlations obtained from meta-analysis were primarily used for ranking, and less as estimates of “true” correlations.

### Average gene expression correlation profile in a dataset collection

An average gene expression correlation profile per query gene in a dataset collection was derived by first selecting different profiles for consistent results between probes, and between datasets, and using maximally one probe per dataset. Since probes for a single gene on a microarray platform can perform differently, also with some probes being inadequate, it was important to select for each microarray dataset the query probe giving the largest average overlap with the correlation results of all other datasets. Profiles were pre-filtered by setting a low threshold (5) for average overlap of the 200 highest ranking genes with all other profiles. Not all datasets gave correlation hits, and this may depend on the quality of specific probes on different platforms, number of samples in a dataset, or type of experiment. In case of smaller dataset collections of separated monocytes, neutrophils, PBMCs and tissues, all available profiles were used to select a single profile for each dataset using largest average overlap with all other profiles, without the pre-filter, to use all datasets in the collection. Next, average correlation profiles were obtained by aggregation of probes per gene in each selected profile using the maximal correlation value with the query gene, and then taking the mean correlation value for all genes in the contributing profiles for ranking, without applying Fisher Z transform, or weighing by dataset sample sizes. Genes present below a threshold number of supporting datasets (6 in case of collections of size 15) were discarded from the combined profile. Since inadequate gene probes, if present, will have higher correlation values with the query (closer to 0), than corresponding adequate gene probes, in case of actual negative correlation, this averaging method is best suited for the positive correlation part of the profiles. To reliably determine the negative correlation end of ranked average gene expression correlation profiles, probes can also be aggregated per gene taking the minimal correlation value. An annotated R script which can be used for multiple gene queries (<u>selectcollections.R</u>), and 2 dataset collections, whole blood in the absence of severe inflammation (n= 15), and whole blood in the presence of severe inflammation (n= 15) are provided to reproduce results of average correlation profiles for single genes using these same collections (Table S1, sheets 1 and 5). This also replicates the results seen using the larger collections (n= 40, and n = 38) (Table S1, sheets 8 and 12).

### Multi-gene signatures

Multiple average gene expression correlation profiles obtained on a given dataset collection, for different co-expressed query genes, can be combined again to give a single robust result for a chosen gene set. In case of using GC-induced gene queries this resulted in multi-gene signatures of GC-driven gene expression. Suitable GC-induced query gene combinations were chosen based on highly similar correlation profiles, which introduced cell type biases in case of blood, or were more balanced for correlation profiles. Ranked gene expression correlation profiles for whole blood GC signatures 1 and 2 with different cell type biases (Table S1) can be reproduced using precalculated results obtained with R script (<u>selectcollections.R</u>) on dataset collections of whole blood in the absence of severe inflammation (n= 15), and whole blood in the presence of severe inflammation (n= 15), using the provided R script (<u>signaturescollections.R</u>). Both R scripts are in folder “code” in GitHub repository (https://github.com/ArthurJohannes/GCblood_repo).

### Relating gene expression correlation profiles

Average gene expression correlation profiles were compared for reciprocal ranking of genes in related profiles to determine the existence of clusters of correlated gene expression by meta-analysis. Using the method described above, reciprocal average correlation values between gene pairs (a,b and b,a) were only identical if identical corresponding gene probes were used, which mostly applies to RNA sequencing data, and when using the same (all) datasets in the dataset collection for both genes. Ranking gene expression correlation values in profiles obtained either on a single dataset or from meta-analysis, results in different reciprocal rankings for gene pairs (rankings for a,b and b,a are different), and asymmetric matrices of gene rankings. An “inside” gene in a large tight co-expression cluster may have higher ranking (closer to 1) in a profile of an “outside” gene than the other way around. This allows easier grouping of the “outside” gene with the large cluster which can be useful, and indicate (marginal) gene induction in case of such clusters resulting from specific gene induction. Initial ordering of rows and columns was guided by hierarchical clustering using the corresponding matrix of reciprocal average correlation coefficients. The order of rows of gene profiles and genes in profiles (columns) was then modified and set to indicate relevant gene groups such as co-expression clusters, and also genes with shared inductions but found outside tight co-expression clusters for the induction, and possibly in other co-expression clusters. The latter groupings were aided by sorting genes present in marker gene profiles according to ranking, and reciprocally, sorting profiles for their ranking of marker genes.

The final chosen setting kept order of genes in rows and columns identical. An annotated R script (<u>reciprocalcollections.R</u>), and precalculated results obtained for 19 genes including sets GC-1 and GC-2, using the R script described earlier (<u>selectcollections.R</u>) replicates similar matrices as shown for larger dataset collections on blood in the absence of severe inflammation (n= 40), and on blood in the presence of severe inflammation (n = 38) (Figure 3A, 3B). Both R scripts are in folder “code” in GitHub repository (https://github.com/ArthurJohannes/GCblood_repo).

### Using gene sets and modules

Different methods exist to summarize expression values of co-expressed genes in a single composite score. Using arithmetic mean expression of all genes in a module is inadequate considering the large differences in relative expression levels between genes. Gene module expression values can be determined for single samples, using the geometrical mean expression of genes in a module (Petri et al., 2019), using eigengenes (Langfelder & Horvath, 2008), single sample GSEA (Barbie et al., 2009), the percentage of upregulated or downregulated genes in a module, compared to a control group, and using normalized expression, such as averaging fold change for individual genes compared to median expression in a control group (Altman et al., 2021; Chaussabel & Baldwin, 2014; Chaussabel et al., 2008), and such as mean of Z-scores for each gene in a set (Petri et al., 2009). While mean of Z-scores, possibly followed by rescaling, might be optimal, to compare module expression between samples in a single experiment, it does not indicate fold change. For a balanced use of GC inducible genes in gene sets GC-1 (n= 7) and GC-2 (n= 9), it was important to account for differences in ratios of maximal and minimal expression for different genes in the set, and also for the presence of microarray platform background levels which varied with specific gene probes, and which also depended on the method used in data processing. Datasets were first converted to linear scale, if necessary. When using gene modules with microarray data, only adequate and identical gene probes were used in case of Affymetrix GPL570, or Illumina GPL10558 microarray platforms, or else gene probes with highest expression levels in the experiment. Gene expression values in a sample were divided by expression range (max-min) of the corresponding gene in the whole dataset, and then multiplied by a constant (sum of gene expression ranges in module divided by number of genes in module) to keep expression values closer to starting values. In case of microarray data, the resultant gene expression values were subtracted with their minimal value to avoid potentially large contribution of background for specific probes. The arithmetic mean was then taken for resultant values of all genes in the module per sample, as a measure of gene module expression. Module expression values were plotted as binned dot plots with boxplot overlay, excluding the boxplot outlier points, on the log2 scale, and using similar Y axis interval sizes to allow for a better comparison between datasets. Annotated R scripts and corresponding datasets are provided in a GitHub repository folder to reproduce results of gene module expression in burn shock (<u>plotmodulesgse77791.R</u>, GSE77791, microarray GPL570 (Plassais et al., 2017)) (Figure S14 C), severe malaria (<u>plotmodulesgse34404.R</u>, GSE34404, microarray GPL10558 (Idaghdour et al., 2012)) (Figure S15 F), and obtain results of single gene, and gene module expression in sepsis (<u>plotgenesgse154918.R</u>, plotmodulesgse154918.R, GSE154918, RNA-seq (Herwanto et al., 2021)) (https://github.com/ArthurJohannes/GCblood_repo/tree/main/plotting).

### Using R and figures preparation

Data were analyzed using R programming language for statistical computing and graphics, in RStudio IDE desktop. The gGPLot2 R package (Wickham, 2016) was used for plotting. For aligning plots, use was made of the R packages grid (Murrell, 2002), gridExtra (Auguie et al., 2017), and package gtable contained in gGPLot2. Package pheatmap (Kolde, 2019) was used for hierarchical clustering of correlation matrices, using Euclidean distance (not directly using correlation dissimilarity as a distance), and clustering method complete, and for custom annotation. R packages xlsx (Dragulescu, 2012), and readxl (Wickham et al., 2019) were used for writing and reading Excel files, or WPS spreadsheet files. Tables were edited in WPS or Excel spreadsheets. Figure PDFs were arranged and edited in Inkscape (Inkscape project, 2020), or created with BioRender.com.

## RESULTS

### Obtaining 2 blood transcriptomic signatures of GC induction

Two different gene expression correlation clusters originating from induction by GC were observed in many blood transcriptomic datasets, one in health, and another in severe inflammation, for example, in health for datasets GSE11761, GSE14642, and GSE57065 (Cazalis et al., 2014; Radom-Aizik et al., 2009a, 2009b), and in critical illness and sepsis for datasets GSE9960 (Tang et al., 2009), and GSE57065 (Figure S1). Exploratory work using programmatically selected datasets from a large collection of 375+ blood transcriptomic datasets indicated that 2 corresponding gene expression correlation signatures could be obtained in meta-analysis: One in the relative absence of severe inflammation (signature 1), the other in the presence of severe inflammation (signature 2). A gene set (n = 227) of candidate GC-regulated genes was derived from high-ranking genes in these earlier signatures of GC-driven gene expression *in vivo* (Table S2). To obtain reproducible, corresponding gene expression correlation signatures, 2 whole blood dataset collections were assembled based on experimental description. The dataset collection in the absence of severe inflammation (n = 40) included health and vaccination, and also excluded diseases where GC usage in treatment is common, such as SLE, RA, and MS. The collection in the presence of severe inflammation (n = 38) mainly included datasets on sepsis (Table S1). Gene expression correlation rankings for all datasets in a single collection were first obtained for a single GC inducible query gene. An averaged correlation ranking for a single query gene used a single best gene probe for each dataset. Results for different suitable query genes were grouped according to cell type bias, and again averaged to obtain multigene expression correlation signatures, namely 2 in the absence of severe inflammation, and 3 signatures in the presence of severe inflammation (Figure 1A and B, Table S1). In case of lymphocyte bias, only a single query gene DDIT4 was used on both collections allowing for a direct comparison of the corresponding signatures (Figure 1B, Table S1). Several query genes such as VSIG4 and ADAMTS2 (both monocyte bias), and OLAH (neutrophil bias) only showed strong GC-driven gene expression correlation in the presence of severe inflammation, whereas other query genes could be used on both collections (Figure 1B). The cell type biased signatures obtained either in the absence of severe inflammation (n =3), or in the presence of severe inflammation (n= 4) are together referred to as GC signatures 1 and 2, respectively. Repeating the procedure on 2 subcollections of 15 datasets each yielded highly similar GC signatures 1 and 2 as shown (Figure 1B, Table S1). Genes PIK3IP1, FBXO32, BTG1, and DUSP2 were typically present in lymphocyte-biased signatures of GC-driven gene expression in whole blood and also in PBMCs in the absence of severe inflammation (Table S1 and S3). Since muscle, skin, and fat represent well known and important tissue targets for GCs, we also applied the procedure to corresponding datasets collections. Distinct but partially overlapping correlation signatures of circadian GC-driven and molecular CLOCK-driven gene expression were readily obtained for these tissues (Table S3). In contrast, CLOCK-driven gene expression was not detected in whole blood and isolated blood cells. High ranking GC signature genes in muscle included transcription factors FOXO3 and FOXO1, and GLUL (encoding Glutamate-Ammonia Ligase) as expected, in skin BCL6 and FOXO1, and in fat, ERRFI1, RHOB, and ITPKC (encoding Inositol-Trisphosphate 3-Kinase C) (Table S3). A good overall signature of GC-driven gene expression in blood and tissues was obtained by ranking genes by frequency of their presence in the first 2000 genes in 16 separate GC signatures (Table S2). More than 1000 candidate GC-regulated genes were indicated by further comparison with *in vitro* upregulated DEGs.

**Figure 1.**
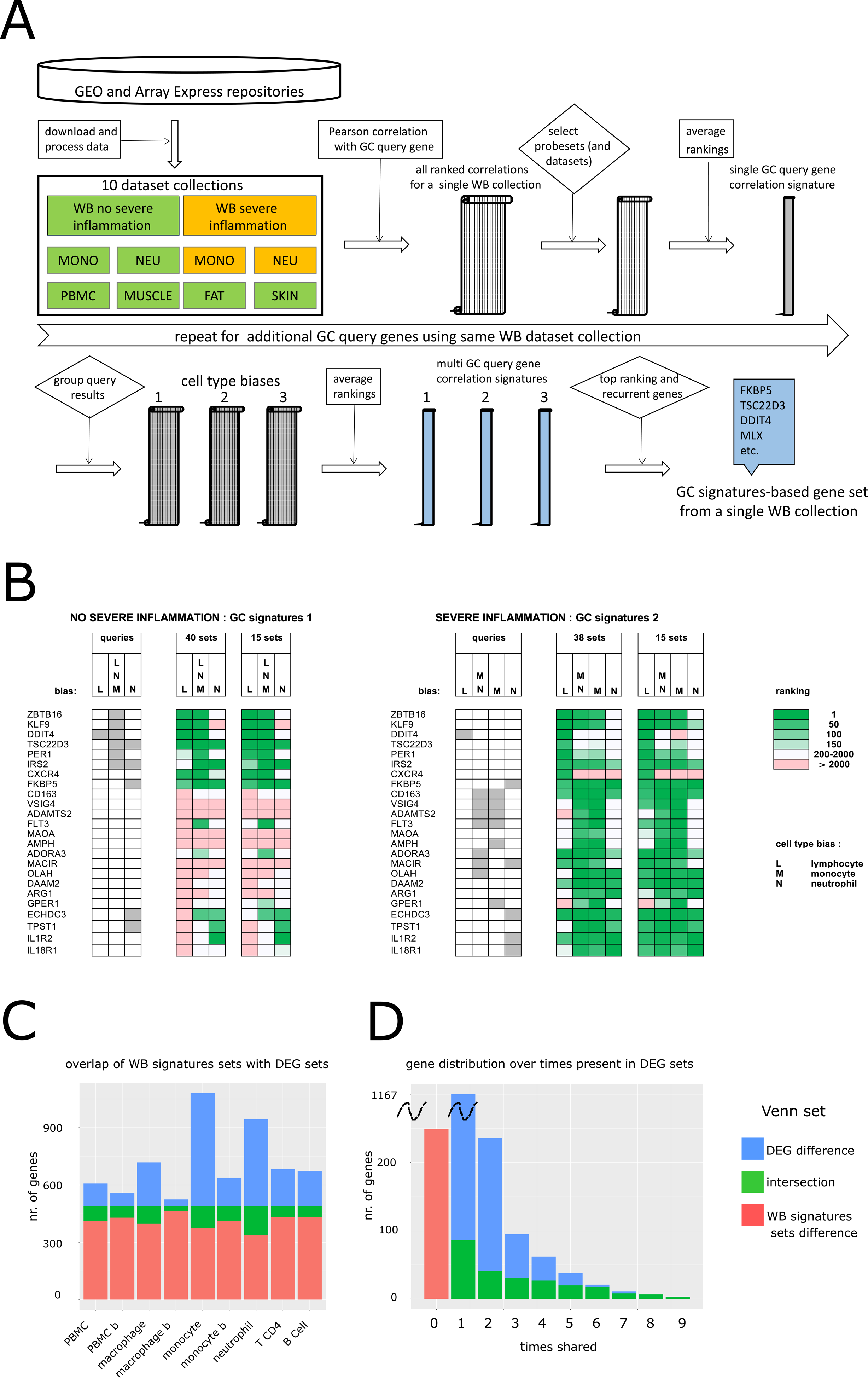
Obtaining GC-driven gene signatures and comparison with *in vitro* DEGs. **A** work diagram for obtaining GC-driven gene expression correlation signatures using defined collections of transcriptomic datasets, as illustrated for a single whole blood collection. Datasets were selected by experimental description. Scrolls symbolize gene lists ordered by Pearson correlation values with query genes. **B** GC-driven gene expression correlation signatures obtained on 2 whole blood dataset collections and subcollections thereof, using appropriate query genes as indicated in grey. **C** Venn diagrams of total gene set derived from whole blood GC signatures 1 and 2, with 9 upregulated DEG sets from GC treatment of immune cells *in vitro*. PBMC (GSE110156, 6h, 1 mM prednisolone (Hu et al., 2018)), PBMC b (GSE33649, 8h, 500 nM dexamethasone (Maranville et al., 2013)), macrophage (GSE61880, time series 1-24h, 100 nM dexamethasone (Jubb et al., 2016)), macrophage b, monocyte (GSE109439, 4h, 1 mM, triamcinolone acetonide (Wang et al., 2019)), monocyte b, neutrophil, CD4 T cell, and B cell (GSE112101, 2h and 6h, 22.7 microM methylprednisolone (Franco et al., 2019)). **D** frequency distribution of all GC-induced DEGs as present in total of 9 DEG sets. Overlap of DEGs with total gene set derived from whole blood GC signatures as indicated.

Among most frequently upregulated genes were negative regulators of MAPK and NF-kB signaling (DUSP1, TSC22D3, NFKBIA), and transcription factors KLF9, ZBTB16, CEBPD, PER1, and ETS2, as expected (Table S2).

### Many high ranking genes in whole blood GC signatures are induced by GC *in vitro*

The signature genes found by correlation search may come from variable differential gene expression due to upregulation by GCs, or more indirectly due to shifts in cell type percentages caused by cortisol in healthy people (GC signature 1), or the appearance of specific cell types in severe inflammation (GC signature 2). Therefore, it was important to determine which signature genes could be observed as DEGs *in vitro*, and at the same time determine the cell types involved at basal and induced gene expression levels. A total gene set of high ranking and/or recurrent genes in whole blood GC signatures 1 and 2 was selected to contain best candidate genes for induction by GC as detailed (Table S2, sheet3). Genes in this set (n = 489) were compared with 9 sets of *in vitro* GC-induced DEGs, taken at a cut-off of 2 fold induction and with unadjusted p-value < 0.05, in different immune cells and PBMC preparations (Figure 1C, 1D, and Table S2). Overlap with different DEG sets was in the range of 5-31 % and higher with myeloid cells than with lymphoid cells in experiment GSE112101 (Franco et al., 2019), suggesting a relatively high contribution of monocyte and neutrophil gene expression to the GC signatures-derived gene set (Figure 1C). Frequent DEGs were well represented in whole blood GC signatures (Figure 1D). Genes FKBP5, SESN1, and TSC22D3 were present in all DEG sets (Table S2). The DEG sets excluded 249 potentially GC-induced genes while 86 genes were detected only once in these experiments. Signature genes that were absent in DEGs included a limited number of NK-expressed genes (PRF1, GZMB, etc.).

These genes originated from the DDIT4 query-derived signature obtained in the absence of severe inflammation, and were likely due to increased NK percentages. The high proportion of genes in the GC signatures-derived gene set upregulated by GC *in vitro* (49 %) indicated that the signatures largely originated from GC gene induction rather than from accompanying shifts in cellular blood count.

### Basal gene expression and induction by GC in different cell types *in vitro*

In a next step, a large number of GC signature genes and different marker genes, used as controls, were directly compared for basal expression levels in separated blood cells, and for their induction by GC *in vitro*. Gene expression was profiled using RNA-seq experiment GSE60424 on separated immune cells from different illnesses, (Linsley et al., 2014) excluding sepsis, RNA-seq experiment GSE112101 which studies GC gene induction *in vitro* in different cell types, including isolated immune cells (Franco et al., 2019), RNA-seq experiment GSE109439 on GC gene induction *in vitro* in monocytes and macrophages (Wang et al., 2019), and microarray experiment GSE100531 on GC gene induction in M1 macrophages (Gharib et al., 2019). Basal level expression data from experiment GSE60424 and GSE112101 were largely similar as expected, although with some differences, e.g. in case of CXCR4, TSC22D3, and FLT3 (Figure S2). A limited number of genes (n= 19) were selected for subsequent expression profiling, including most genes used as queries to obtain whole blood GC signatures 1 and 2, as well as additional genes used in 2 gene sets GC-1 and GC-2 drawn up for possible biomarker use (see below) (Figure 2A). Gene ZBTB16 was preferentially expressed in B and NK cells, IRS2 was highly expressed in neutrophils, and KLF9 had low basal expression in neutrophils in both experiments GSE112101 (Franco et al., 2019) and GSE60424 (Linsley et al., 2014) (Figure 2B, and Figure S2). Genes CD163, VSIG4, and ADAMTS2 were specifically expressed in monocytes, FLT3 and MAOA preferentially in monocytes, ADORA3 in both monocytes and neutrophils, while OLAH, DAAM2, and ARG1 were preferentially expressed in neutrophils (Figure 2B). Basal expression of ADAMTS2 and FLT3 was higher in monocytes than in monocyte-derived macrophages, while expression of CD163, VSIG4, MAOA, and ADORA3 genes was higher in macrophages instead. *In vitro* upregulation by GC in different separated immune cells and in macrophages was visible at least once for these 19 genes, sometimes even in cell separations with very low/absent basal expression, as in case of CD163 in B cells and VSIG4 in T cells, probably due to small amounts of contaminating myeloid cells (Figure 2B). Gene ARG1 was upregulated by GC in neutrophils (Figure 2B).

**Figure 2.**
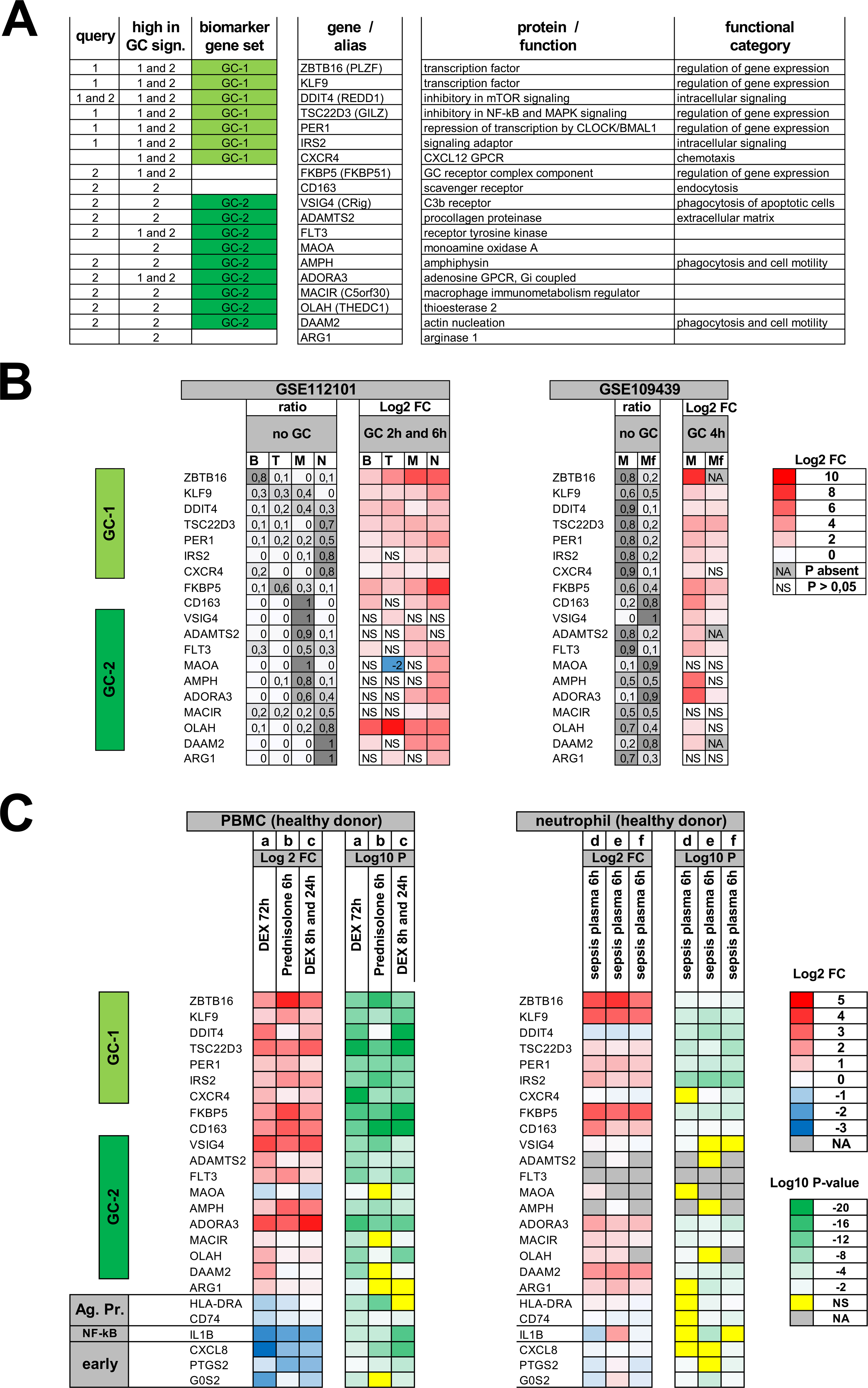
In vitro gene induction by GC. Overview of selected genes used for expression profiling, and *in vitro* induction by GC in PBMCs and immune cells. **A** Overview of 19 genes selected from GC-signatures. Query: genes used as expression correlation queries to obtain GC signatures 1 and 2 from 2 whole blood dataset collections. High in GC-sign.: high ranking of gene in GC signatures. Biomarker set: genes selected in GC-1 and GC-2 biomarker sets. **B** Induction in isolated immune cells (GSE112101, (Franco et al., 2019)). Ratio: ratio of total expression at time 0h comparing 4 types of blood cells (B, B cells; T, CD4 T cells; M, monocytes; N, neutrophils). Log2 FC: fold change after GC treatment; methylprednisolone 22.7 microM, 2h, and 6h versus vehicle control (pairwise for 4 donors). NS: pairwise p-value > 0.05 (left panel). Induction in monocytes (M), and macrophages (Mf) (GSE109439, (Wang et al., 2019)). Ratio: ratio of total expression at time 0h in monocytes and macrophages. Log2 FC: fold change after GC treatment; triamcinolone 1 mM, 4h versus vehicle control (right panel). **C** Induction by GC in PBMCs; GSE217320, (Seah et al., 2022) (a), GSE110156, (Hu et al., 2018) (b), GSE33649, (Maranville et al., 2013) (c). Log 2FC: treatment versus vehicle controls; dexamethasone 2.5nM, 5nM, 50nM combined, 72h (a); prednisolone 1mM 6h (b); dexamethasone 0.5 mM 8h, and 24h (c) (left panel). Induction by sepsis plasma in isolated neutrophils; GSE49755 (d), GSE49756 (e), GSE49757 (f), (Khaenam et al., 2014) (right panel). Unadjusted p-values as indicated. Marker genes for antigen presentation (Ag. Pr.), NF- kB-dependent expression (NF-kB), and early gene expression (early) are also included.

### Frequent GC activity in septic plasma in neutrophil reporter assays

GC treatment *in vitro* will result in many upregulated and downregulated genes, also depending on duration of treatment. The 19 selected genes were upregulated by *in vitro* GC treatment of PBMCs in 3 experiments from 6h to 72h (Hu et al., 2018; Maranville et al., 2013; Seah et al., 2022), with the exception of MAOA (Figure 2C left panel). Downregulation of HLA-DR marker gene expression by GC was visible, in agreement with the known transcriptional effect of GC on monocytes *in vitro* (Tulzo et al., 2004). Inhibition by GC of early gene activation in PBMCs (Figure 2C left panel), occurring *in vitro,* agreed with inhibition of pro-inflammatory signal transduction by GC, involving MKP-1 (DUSP1), and GILZ (TSC22D3), as described for monocytes (Ehrchen et al., 2019). Khaenam et al., (2014) reported a set of 30 gene transcripts responsive to septic plasma in neutrophil reporter assays, which they used to predict sepsis severity. This set contained 9 genes also prominently present in the GC signatures reported here, including well-known GC inducible genes such as KLF9 and FKBP5. Many other GC-regulated genes were also induced by sepsis plasma, including 13 out of the 19 pre-selected genes used here for expression profiling (Figure 2C right panel), indicating relatively high concentrations of GC compared to control plasma. Strong induction could be observed at a high frequency of approximately 50 in 73 unique sepsis plasma, above the maximal induction level seen in the healthy control plasma. Since potential GC usage was not reported for these experiments (Khaenam et al., 2014), it could not be determined whether this frequent GC activity might also be endogenous, or just always derived from exogenous GC in treatment. Sepsis plasma from neutrophil reporter assay experiment GSE49756 also showed additional immunomodulatory activities corresponding with induction by interferon gamma, as well as NF-kB-dependent induction, as shown by upregulation of marker gene IL1B (Figure 2C right panel).

### Circadian GC, and GC in severe inflammation, drive different patterns of gene expression in blood, monocytes, and neutrophils

To examine differential expression correlation of GC signature genes in health and in severe inflammation more precisely, we used reciprocal ranking of genes in expression correlation profiles, directly comparing the 19 preselected genes. Separate collections of monocyte and neutrophil transcriptomic datasets were assembled, also based on the absence or presence of severe inflammation, according to provided metadata and experimental description.

Collections of (sub)datasets in the absence of severe inflammation included only healthy controls in case of both monocytes (n= 8), and neutrophils (n= 6), while collections in the presence of severe inflammation included datasets mainly on sepsis for either monocytes (n= 11) or neutrophils (n= 10) (Table S3). Different correlation patterns were present dependent on the presence or absence of severe inflammation in blood, and also in isolated monocytes and neutrophils (Figure 3). In blood, in the absence of severe inflammation, expression of genes MAOA, ADORA3, and CD163 was more correlated with cell type specific expression (reticulocytes, eosinophils, and classical monocytes, respectively), while genes ADAMTS2, and AMPH, failed to yield good average gene expression correlation profiles, due to detection limits (Figure 3A). In the presence of severe inflammation, expression was more correlated, with the exception of the gene encoding CXCR4 (Figure 3B). From this analysis, using additional gene expression correlation, and differential expression data, we selected GC-1 (ZBTB16, KLF9, DDIT4, TSC22D3, PER1, IRS2, CXCR4), and GC-2 (VSIG4, ADAMTS2, FLT3, MAOA, AMPH, ADORA3, MACIR, OLAH, DAAM2) gene sets for possible use as whole blood transcriptomic biomarkers in health, and severe inflammation, respectively, as detailed (Figure S3, and Figure S4). In the absence of severe inflammation, clustering was best visible for set GC-1, and also FKBP5, which was tighter in monocytes than in blood (Figure 3A, 3C), likely due to a lower cellular complexity. Datasets on isolated monocytes and neutrophils often showed evidence of contaminating cells, confounding gene expression correlation results to some extent. ARG1 expression correlated with other high ranking GC signature genes in neutrophils in severe inflammation, but not in monocytes (Figure 3D, 3F). High expression correlation between VSIG4 and ADAMTS2 was also visible for neutrophils in severe inflammation (Figure 3F), likely due to monocyte contamination. A comparison of RNA-seq datasets with circadian sampling in health (Braun et al., 2018), and in bacterial infection and sepsis (Herwanto et al., 2021) indicated tighter clustering for genes in set GC-2 in the latter (Figure 3G, 3H). Large differences in relative mRNA abundance (appr. > 1000 x) existed between the 19 genes based on TPM, and normalized count values, with TSC22D3, and CXCR4 mRNA being more abundant, and in severe inflammation, also enhanced expression of IRS2, FKBP5, CD163, and ARG1 mRNA was found (Figure 3G, 3H).

**Figure 3.**
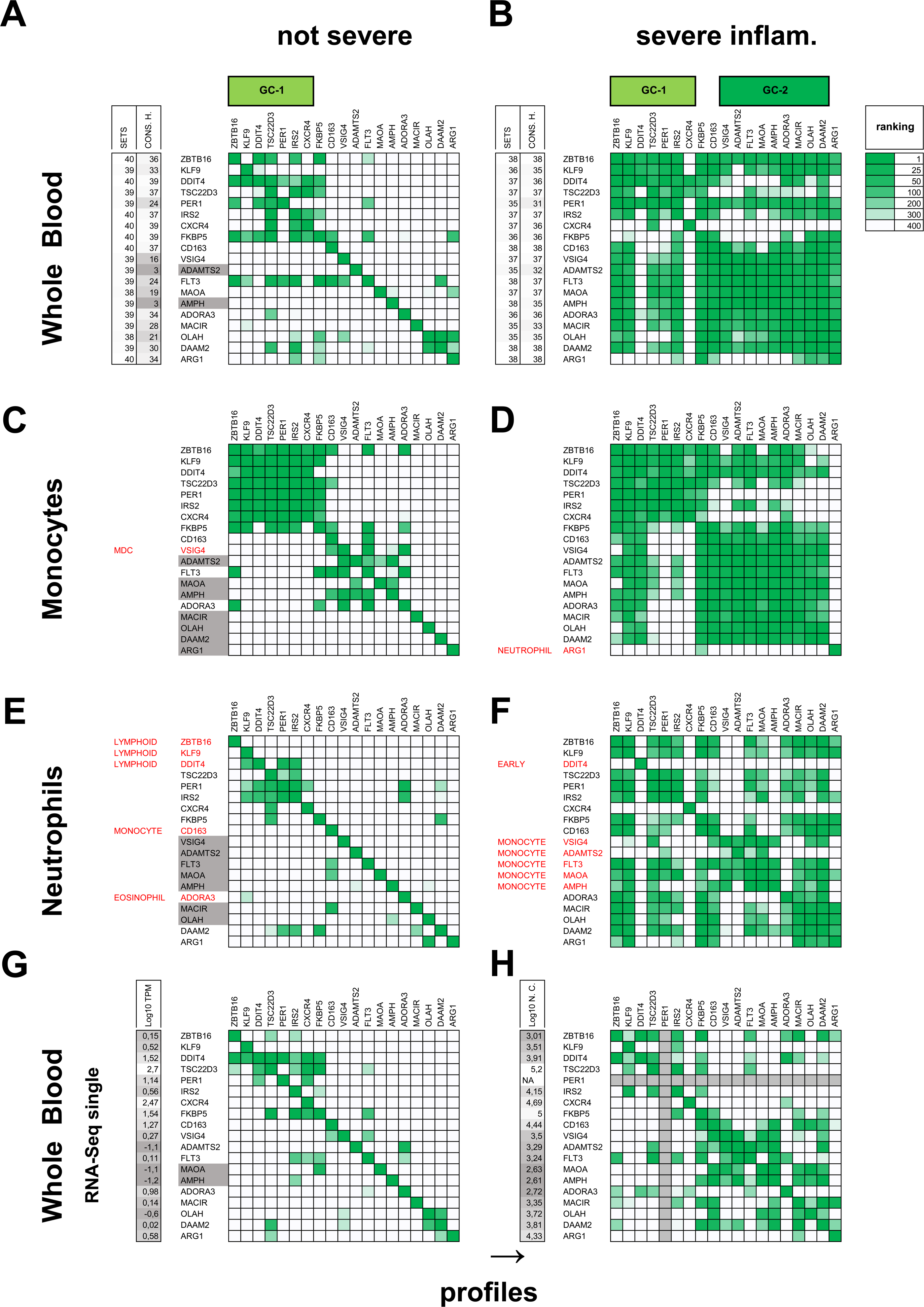
Differential expression correlation of genes induced by GC in health and severe inflammation. Reciprocal ranking in gene expression correlation profiles for 6 dataset collections and 2 separate RNA-seq datasets. **A-B** profiles for whole blood. Number of datasets with gene (SETS), and consistent gene expression correlation hits (CONS. H.), as indicated. Genes selected as biomarker sets GC-1, and GC-2 as shown. **A** whole blood in the absence of severe inflammation (40 sets in collection). **B** whole blood in the presence of severe inflammation (38 sets in collection). **C-F** profiles for myeloid cells. Genes in grey; expression below detection limit in all or most datasets, genes in red; profile originating from other contaminating cell types, or *in vitro* activation. **C** monocytes in the absence of severe inflammation (8 (sub)sets in collection). **D** monocytes in the presence of severe inflammation (11 (sub)sets in collection). **E** neutrophils / granulocytes in the absence of severe inflammation (6 (sub)sets in collection). **F** neutrophils / granulocytes in the presence of severe inflammation (10 (sub)sets in collection). **G** whole blood, healthy, circadian sampling (GSE113883, (Braun et al., 2018)), excluding one outlier individual no. 14. Genes MAOA and AMPH in grey; at detection limit. Log10 TPM: log of mean TPM values in all samples. **H** whole blood, sepsis, and uncomplicated infections (GSE154918, (Herwanto et al., 2021)). Log10 N. C.; log of mean normalized counts in all samples for ill patients. Gene rankings in profiles color-coded in green from 1 (high) to 400 (low). Gene expression correlation profiles for each of 19 genes in Figure 2A shown in rows.

### Gene induction in vivo by endogenous GC and experimental GC treatment in health

The presence of 2 signatures of GC-driven gene expression in blood in health and in severe inflamamtion might result from different relative induction levels of GC target genes, primarily depending on the concentration and duration of the GC stimulus, or alternatively, might be more conditional on the absence or presence of severe inflammation, and the accompanying cell types. To first address these alternative possibilities, GC-upregulated gene expression *in vivo* was compared using 3 informative PBMC gene expression datasets.

Experiment GDS3704 (Bouwens et al., 2010), using PBMC samples taken 6h apart, did not show large shifts in cell percentages between time points, as indicated by similar NK and monocyte marker gene expression levels (Figure 4A). As a consequence, GC signature genes represented a high proportion of upregulated DEGs at the earlier timepoint (Table S4 sheet 1). Both monocyte and lymphoid cell expressed gene clusters were visible among GC signature DEGs (Figure S5), with tight clustering of set GC-1 genes, and also FKBP5 (Figure 4C).

**Figure 4.**
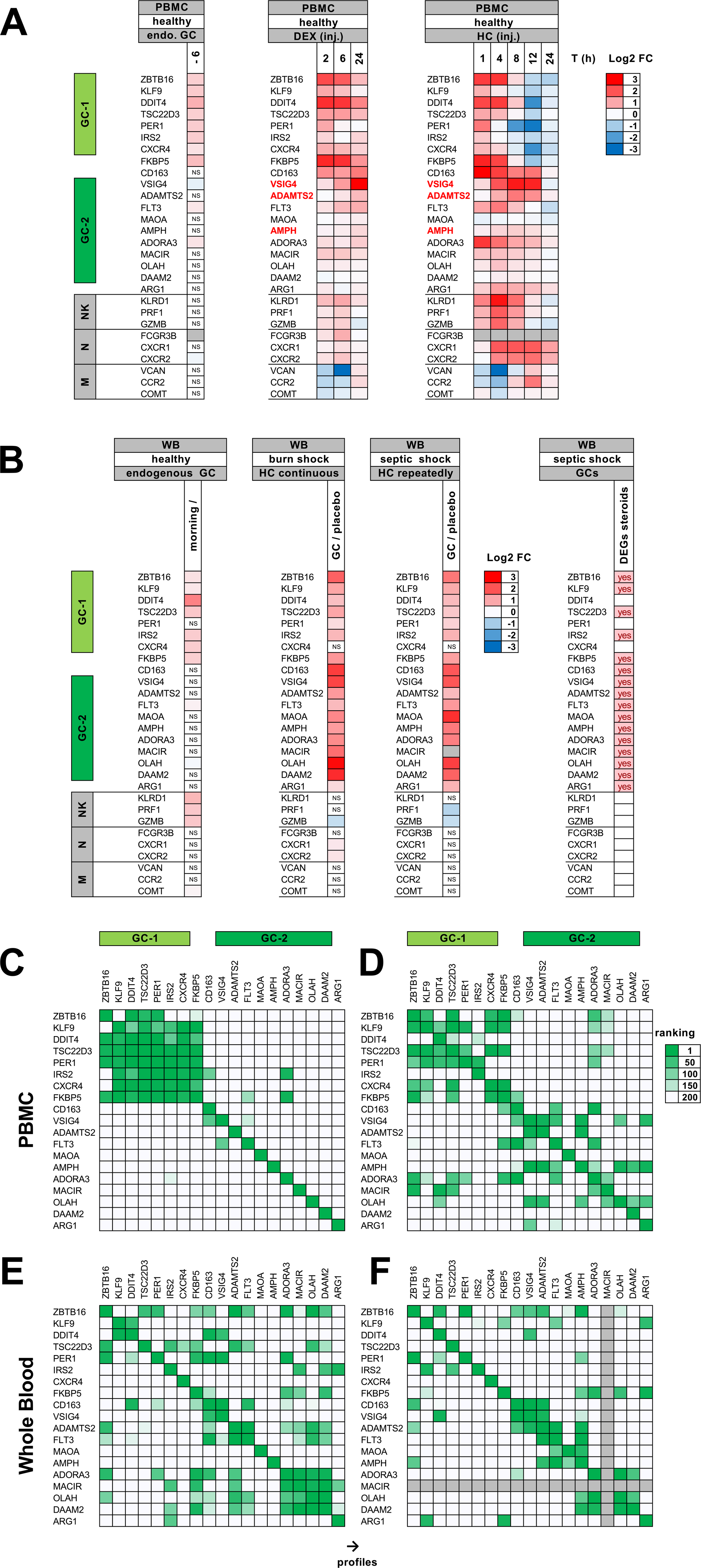
In vivo gene induction by GC. Gene expression changes due to endogenous and exogenous GC in health, and in septic and burn shock. **A** DEGs in PBMCs by endogenous GC, and by experimental GC treatment in health. DEGs in 6h time difference, due to endogenous, circadian cortisol (GDS3704, (Bouwens et al., 2010)), (left panel). DEGs after dexamethasone (DEX) injection, compared with time 0h (GDS3864, (Carlet et al., 2010)) (middle panel). DEGs after hydrocortisone (HC) injection (GSE67255, (Olnes et al., 2016)). Doses 50 and 250 mg combined, and compared with time 0h (right panel). Gene names in red: tight correlation clustering observed with gene VSIG4 in dataset. Selected genes as also shown in Figure 2A, biomarker sets GC-1, and GC-2 as indicated, with added marker genes for expression in NK cells (NK), neutrophils (N), and monocytes (M). **B** DEGs in whole blood by endogenous GC in health, and by exogenous GC in burn shock and septic shock. DEGs between morning compared with rest of day, due to endogenous, circadian cortisol (GDS2767, (Baty et al., 2006)) (left panel). DEGs due to hydrocortisone treatment in burn shock (GSE77791, (Plassais et al., 2017)), comparing 24h and 120h continuous GC treatment, with 24h and 120h placebo, and in septic shock (GSE106878, (Kolte, 2020)), comparing 24h GC treatment with 24h placebo (middle panels). DEGs due to GC treatment in septic shock as reported by Wong et al. (2014) (right panel). Marker genes added for expression in NK cells (NK), neutrophils (N), and monocytes (M). **C-F** reciprocal gene expression ranking in profiles from 4 single data (sub) sets. **C** PBMCs in health, effect of endogenous GC (GDS3704, (Bouwens et al., 2010)), **D** PBMCs in health, effect of exogenous GC (GSE67255, (Olnes et al., 2016)), **E** whole blood, burn shock, subset no GC treatment, endogenous GC (GSE77791, (Plassais et al., 2017)). **F** whole blood, septic shock, subset no GC treatment, endogenous GC (GSE106878, (Kolte, 2020)). Gene rankings in profiles color-coded in green from 1 (high) to 200 (low). Gene expression correlation profiles for each of 19 genes in Figure 2A shown in rows.

Genes VSIG4, ADAMTS2, CD163, and AMPH were not detectably upregulated, while other monocyte expressed GC-induced genes, such as FLT3, ALOX15B, GPER1, and MARVELD1 could be detected as DEGs in this experiment (Figure 4A, and Table S4 sheet 1). In contrast, experimental *in vivo* GC treatment in health (Carlet et al., 2010; Olnes et al., 2016) caused large shifts in cell percentages in PBMCs with increased NK and decreased monocytes at earlier time points (Figure 4A), as described (Olnes et al., 2016). In these experiments, VSIG4, ADAMTS2, and AMPH were co-upregulated upon *in vivo* GC treatment (Figure 4A, 4D), indicating that detectable induction of these genes in PBMCs was not strictly conditional to severe inflammation, and more a consequence of GC stimulus strength above circadian levels.

### Gene induction *in vivo* by endogenous and therapeutic GC in sepsis and burn shock

Recent blood (cell) transcriptomic data, taken together, describe a large number of GC inducible genes in blood in therapy with GC (see introduction). In case of septic shock, 204 genes expressed in blood were reported to be modulated by GC treatment (Wong et al., 2014), and 175 genes in case of burn shock (Plassais et al., 2017). However, likely GC target genes in upregulated DEGs were not identified by these studies, beyond a limited number (n = about 5) using IPA (Ingenuity pathway analysis) (Plassais et al., 2017). Another dataset GSE106878 on GC treatment of septic shock, was published by Kolte and co-workers (2019), but without full listing or identification of GC upregulated DEGs (Kolte, 2020). Here we compared upregulated DEGs from these 3 whole blood microarray datasets on GC treatment in septic and burn shock, and checked their *in vitro* upregulation by GC in monocytes and neutrophils (Franco et al., 2019), and their ranking in GC signatures 2, and signatures of GC-driven gene expression from isolated neutrophils and monocytes in severe inflammation. Extensive overlap was found, thus extending the number of likely GC-induced genes (n > 150) among upregulated DEGs, including set GC-1 and set GC-2 genes, with the exception of CXCR4 in set GC-1 (Table S4). In GC-treated septic and burn shock, shifts in cell type percentages caused by GC treatment were limited (Figure 4B), likely resulting in a large proportion of GC inducible genes in the upregulated DEGs. As in case of PBMCs in health (Figure 4A), set GC-2 was not useful to monitor circadian GC action in whole blood (Figure 4B). A time course experiment of whole blood gene expression in health (GDS2767 (Baty et al., 2006)), showed upregulation of several GC-1 set genes including CXCR4, but not GC-2 set genes. An increase in NK cell percentages was found in the morning (Figure 4B). To determine whether the presence of GC signature 2 in severe inflammation would always be related exclusively to GC treatment, we examined gene expression correlation patterns in untreated patient groups. Correlation patterns in GC-untreated burn and septic shock data were also indicative for GC signature 2, for example by high ranking of GC-2 set genes in ZBTB16, ADAMTS2, CD163, and ADORA3 profiles (Figure 4E, 4F). This analysis showed a specific effect by endogenous GC before treatment, on blood gene expression in severe inflammation, which might depend on the presence of an increased GC stimulus compared to circadian GC in health.

### Using gene set GC-1 as a transcriptional biomarker for circadian cortisol

Major sources of intra-individual variability in cellular blood count in health are circadian rhythm (Ackermann et al., 2012), and exercise (Gustafson et al., 2017). As a result, relative gene expression changed in a circadian fashion, which was in close relation to circadian changes in differential (relative) blood counts. Well visible circadian expression in longitudinal respiratory viral challenge experiment GSE73072 (Liu et al., 2016) was used as a co-selection criterion for set GC-1 (Figure S3B), to minimize the confounding effect of circadian change in cell type percentages. This experiment sampled whole blood, during several days, mostly at 3 time points per twenty-four hours for a larger number of individuals, using a microarray platform. C1QA gene expression in non-classical / intermediate monocytes was upregulated during inflammation in influenza, compared to marker genes used for neutrophils (Figure 5, row 1 panels) and classical monocytes (Figure 5, row 2 panels).

**Figure 5.**
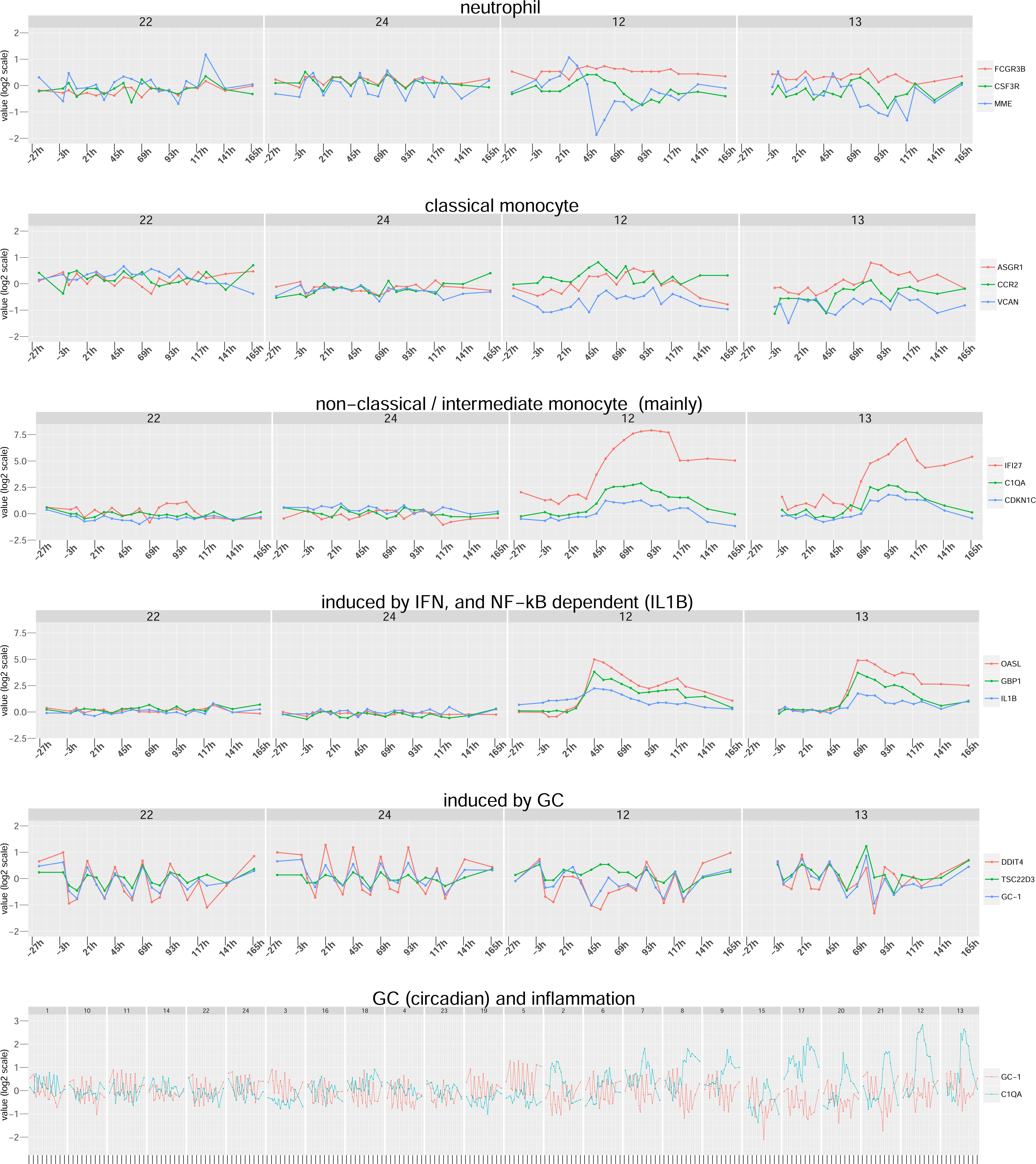
Circadian cortisol action detected using biomarker gene set GC-1. Gene expression in time series of experimental respiratory viral infection, showing marker genes for cell type and inflammation, GC inducible genes DDIT4 and TSC22D3, and set GC-1. Panels rows 1 -5) Gene expression in 2 nonresponding (Nos 22 and 24) and 2 responding (Nos 12 and 13) individuals infected with H1N1 from experiment GSE73072 (Liu et al., 2016). X-axis, time (h) relative to viral challenge at 0h. Y-axis, expression value at log 2 scale. Row 1) marker genes for neutrophil-specific expression. Row 2) marker genes for classical monocyte-specific expression. Row 3) marker genes for non-classical / intermediate monocyte-specific expression. Row 4) marker genes for ongoing inflammation. Row 5) genes DDIT4 and TSC22D3, and combined value for 7 genes in set GC-1. Row 6) Gene expression in all 24 individuals infected with H1N1 from experiment GSE73072, nonresponding (n= 13), and responding (n= 11), showing C1QA, used as marker gene for inflammation, and set GC-1. X-axis, 24h interval ticks in phase with maximal expression of GC-1. Y-axis, expression value at log 2 scale.

Gene induction also marked ongoing inflammation, for example, in case of IL1B, interferon alpha inducible genes OASL and IFI27, and interferon gamma inducible GBP1 (Figure 5). Expression of all 7 genes in set GC-1 was visibly circadian (Figure S3B), with highest expression at early day as shown for DDIT4 (mostly expressed in NK cells), and TSC22D3 (mostly neutrophil) (Figure 5). Several GC signature 1 genes were upregulated during inflammation (eg SOCS1, NFIL3, CLEC4E, PELI1, TNFAIP3), but not genes selected in set GC-1 (Figure S3B). A composite value of gene expression in the set (see methods) was used to visualize circadian cortisol action in health, which also continued during mild inflammation, but from a lower basal level. Some variability in mean and range of DDIT4, TSC22D3, and set GC-1 expression between healthy individuals was apparent, which may relate to small individual differences in circadian cortisol phase, longer lasting individual differences in cellular percentages (as a confounder) or to actual individual differences in strength of GC induction (Figure 5). In conclusion, gene set GC-1 was suitable for longitudinal monitoring of circadian GC activity in whole blood, and was not induced by systemic mild inflammation caused by uncomplicated respiratory viral infection.

### Specific gene induction in severe inflammation compared to mild inflammation

GC inducible gene expression was upregulated during severe inflammation but not during mild inflammation, as in case of set GC-1 genes which retained circadian expression in experimental respiratory viral infection. For a better view of GC signature gene expression in severe inflammation, it was important to first differentiate gene expression in mild and severe inflammation more generally. Therefore, datasets on mild respiratory viral infection (Zhai et al., 2015), and sepsis (Burnham et al., 2017), were directly compared for gene expression correlation, and differential expression with healthy controls, using identical Illumina gene probes. Longitudinal data on mild seasonal respiratory viral infection, agreed well with experimental viral challenge (GSE73072, (Liu et al., 2016), as visible for interferon alpha inducible genes OASL, and IFI27, still upregulated at day 6 after first symptoms, especially in influenza, as reported by Zhai et al., (2015), (Figure 6B). Differential expression correlation between disease days 0-6 in mild respiratory viral infection, and sepsis was determined for marker genes for different cell types and specific inductions. Gene expression correlation clusters originating in inflammation were conserved to different degrees in respiratory viral infection and sepsis. More conserved clustering included marker genes for induction by interferon alpha and gamma, genes CASP1 and CASP4, and other genes indicated in red (Figure 6A). Different or new clustering in severe inflammation included GYG1 with CEACAM1 and MMP8 in neutrophils, and C1QA/B/C genes with PPARG and VSIG4 in monocytes, as indicated in black (Figure 6A right panel). STAT3-dependent inductions of SOCS3, and SBNO2 were associated with inflammation, and development, respectively.

**Figure 6.**
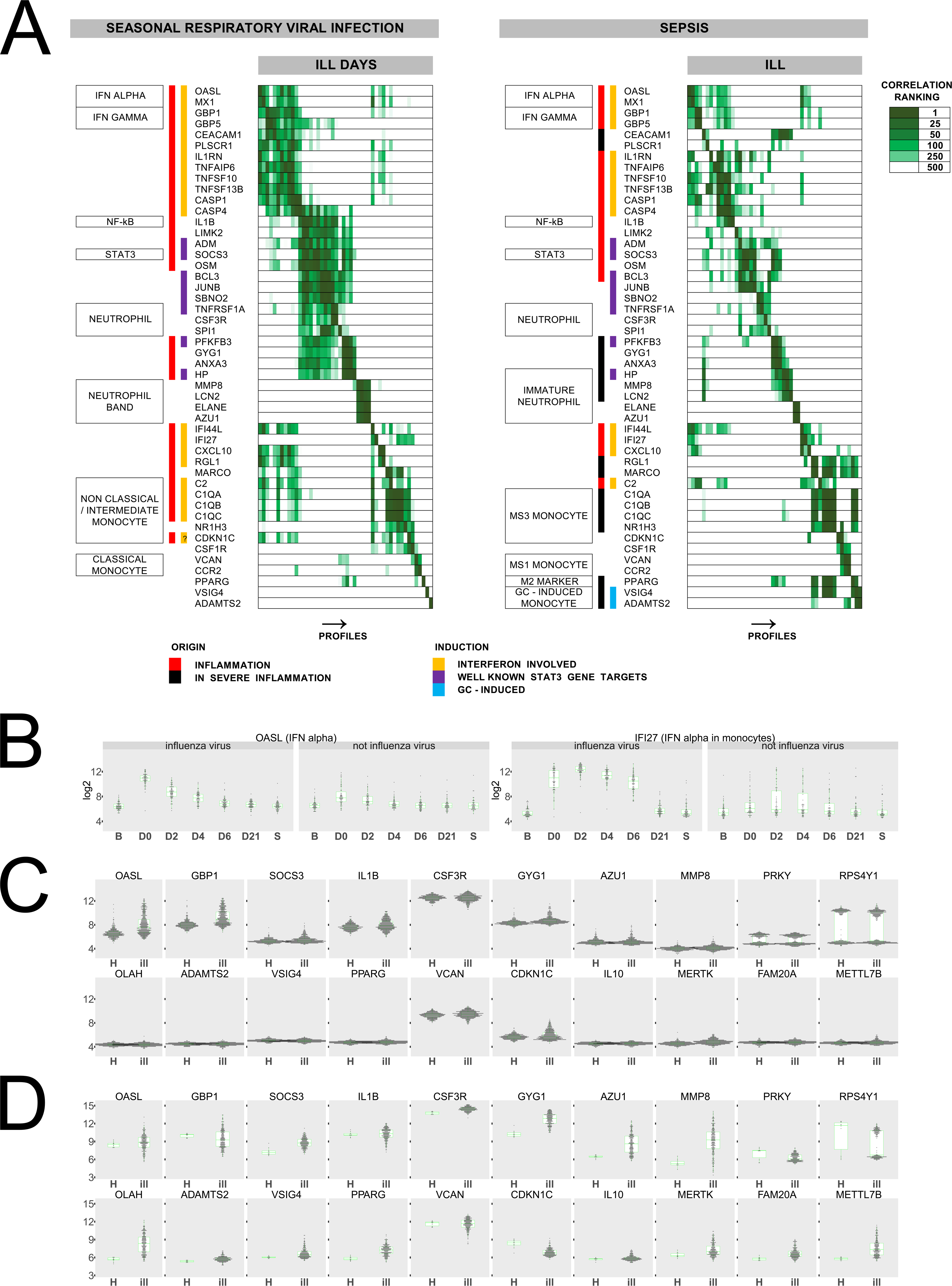
Specific gene expression in mild and severe inflammation. Differential correlation and differential gene expression in whole blood in mild and severe inflammation. **A**) reciprocal gene ranking in 48 gene expression correlation profiles (rows) for mild seasonal respiratory viral infection (GSE68310, (Zhai et al., 2015), subset days 0, 2, 4, and 6 after reporting ill), and for sepsis (E-MTAB-5273, (Burnham et al., 2017), subset sepsis and bacterial infection). Linear scale was used in obtaining correlation profiles, only shared gene probes for both datasets, and in case of 48 query genes, only single adequate gene probes. Genes profiles were put in same order, for easier comparison. Gene rankings in profiles shaded in green (high) to white (low). Marker genes for cell types and inductions as shown, including monocyte states MS1 and MS3 according to Reyes et al. (2020), PPARG (M2 monocyte / macrophage), IL1B (NF-kB-dependent expression), and SOCS3 (STAT3- dependent expression). Origin of different correlation clusters in inflammation and in severe inflammation as indicated (origin, see main text). Genes (co)-induced by interferon, genes likely dependent on STAT3, and genes induced by GC as indicated (induction). **B**) longitudinal expression of interferon alpha inducible genes OASL and IFI27 in dataset GSE68310 on prospective, seasonal respiratory viral infection, either influenza, or not influenza (B; baseline, D0, D2, D4, D6, D21; days after reporting ill, S; spring). **C**, **D**) gene expression distributions in ill and healthy sample groups for a selection of genes used in correlation comparison, and several additional relevant genes (e.g. IL10 en FAM20A). **C**) gene expression in dataset GSE68310 on prospective, seasonal respiratory viral infection, using ill sample groups; 0, 2, 4, and 6 days (ill) and healthy sample groups; baseline, 21 days, and spring (H). **D**) gene expression in dataset E-MTAB-5273 on sepsis in sample groups sepsis or bacterial infection (ill), and healthy (H). **B**, **C**, **D**) Y-axis values at log-2 scale, with interval of 12 log-2 units for each panel. A partial BG subtraction of 150 units at linear scale was performed for GSE68310. Genes PRKY and RPS4Y were used as 2 controls showing signal in male samples, and BG noise level and distribution in female samples. Shown are box plots with added binned dot plots. Identical gene probes from platform GPL10558 used in C and D.

Gene expression clusters corresponding to classical monocytes, and non-classical / intermediate monocytes were visible in mild inflammation, and related clusters corresponding to MS1 and MS3 monocyte states (Reyes et al., 2020) in sepsis (Figure 6A). Combined correlation and differential expression analysis of genes AZU1 and MMP8 indicated that neutrophils with banded nuclei were not increased during mild inflammation, while these immature neutrophils were increased in severe inflammation (Figure 6A, 6C, 6D), consistent with emergency granulopoiesis occurring in sepsis (Kwok et al., 2023). Gene GYG1 encoding glycogenin, used as a marker for glucose metabolism mainly in neutrophils, was more upregulated at a higher frequency in sepsis than in mild inflammation (Figure 6C, 6D). Other genes, highly upregulated in severe inflammation, but not in mild inflammation, included GC signature 2 genes OLAH (mainly in neutrophils), ADAMTS2, and VSIG4 (both in monocytes), as well as genes not regulated by GCs such as FAM20A, and METTL7B (Figure 6C, 6D). Inductions involving interferon were also visible in sepsis, but from a relatively lower basal level, possibly due to large shifts in global gene expression.

### Gene induction by GC in myeloid cells in health and during severe inflammation

In addition to GC signatures 1 and 2, signatures of GC-driven gene expression were also obtained for monocytes, and neutrophils, in each case distinguishing between the presence or absence of severe inflammation according to description (Table S1, Table S3). These signatures predicted wide ranging effects of GCs on cell biology. Based on gene functional categories, this included modulation of phagocytosis and cell motility, modulation of secretory pathway activity, increased sphingolipid synthesis, and uptake of apoptotic cells (efferocytosis) by monocytes during severe inflammation (Figure S6). Specific gene sets for monocytes and neutrophils were derived by combining relevant GC signatures (Table S2) increasing the total number (n = 625) of candidate GC-regulated genes recovered from blood and separated cells (Table S2). Comparing with *in vitro* data showed 50 % of these genes being upregulated by GC at least once in 9 experiments (Table S2). To relate differences in GC-regulated gene expression to inflammation status, we used the monocyte and neutrophil dataset collections, that were aided by data on *in vitro* inducibility in myeloid cells by interferons and interleukins (GSE190594; (Green et al., 2023), GSE131990; (Devlin et al., 2020), GSE146438; (Gorby et al., 2020)), and *in vivo* differential expression in health, interferon treatment in MS, and sepsis (GSE60424; (Linsley et al., 2014), GSE133822; (Washburn et al., 2019), GSE123729; (Coulibaly et al., 2019)). Isolation of monocyte and neutrophils from whole blood generally resulted in early gene activation, as was evident from increased expression of immediate early genes, and the presence of corresponding gene expression correlation clusters (Figure S7, S8, S9, S10). Whole blood gene expression data were used instead to determine *in vivo* expression of such genes in neutrophils, in case of mainly neutrophil-derived gene expression clustering. Gene expression originating from contaminating cells was identified by using suitable marker genes (Figure S7, S8, S9, S10). An overview shows the presence of different myeloid cell types and gene inductions as seen in homeostasis, inflammation, and in severe inflammation (Figure 7). Gene expression correlation clusters corresponding to neutrophils with banded nuclei and (meta)myelocytes were visible in collections of control neutrophils (Figure S9), and neutrophils in severe inflammation (Figure S10), respectively. Intermediate / non-classical monocytes were present in the absence of severe inflammation (Figure S7). A gene expression correlation cluster of C1QA/B/C genes, OLFML2B, SLCO2B1, VSIG4, MSR1, MRC1 (encoding M2 macrophage marker CD206), and LYVE1 (macrophage marker in mononuclear phagocytes), in severe inflammation, indicated the presence of M2 macrophage-like monocytes (Figure S8). Gene induction involving interferons alpha and gamma was well visible in dataset collections of control neutrophils and monocytes (Figure S7, S9), likely due to a low frequency of samples with mild inflammation occurring in the control groups. Gene induction in monocytes and neutrophils during mild inflammation, not involving interferon, included marker gene IL1B (NF-kB-dependent, likely induced by cytokines IL1B and/or TNF), and other genes of which several with known STAT3-dependent induction (eg marker gene SOCS3, likely induced by cytokines G-CSF and/or IL6) (Figure S7, S9). Inductions involving interferon remained present during severe inflammation, for example in case of OASL (IFN alpha), GBP2 (IFN gamma), and CASP1 (possibly both IFN-induced and NF-kB-dependent) (Figure S8, S10).

**Figure 7.**
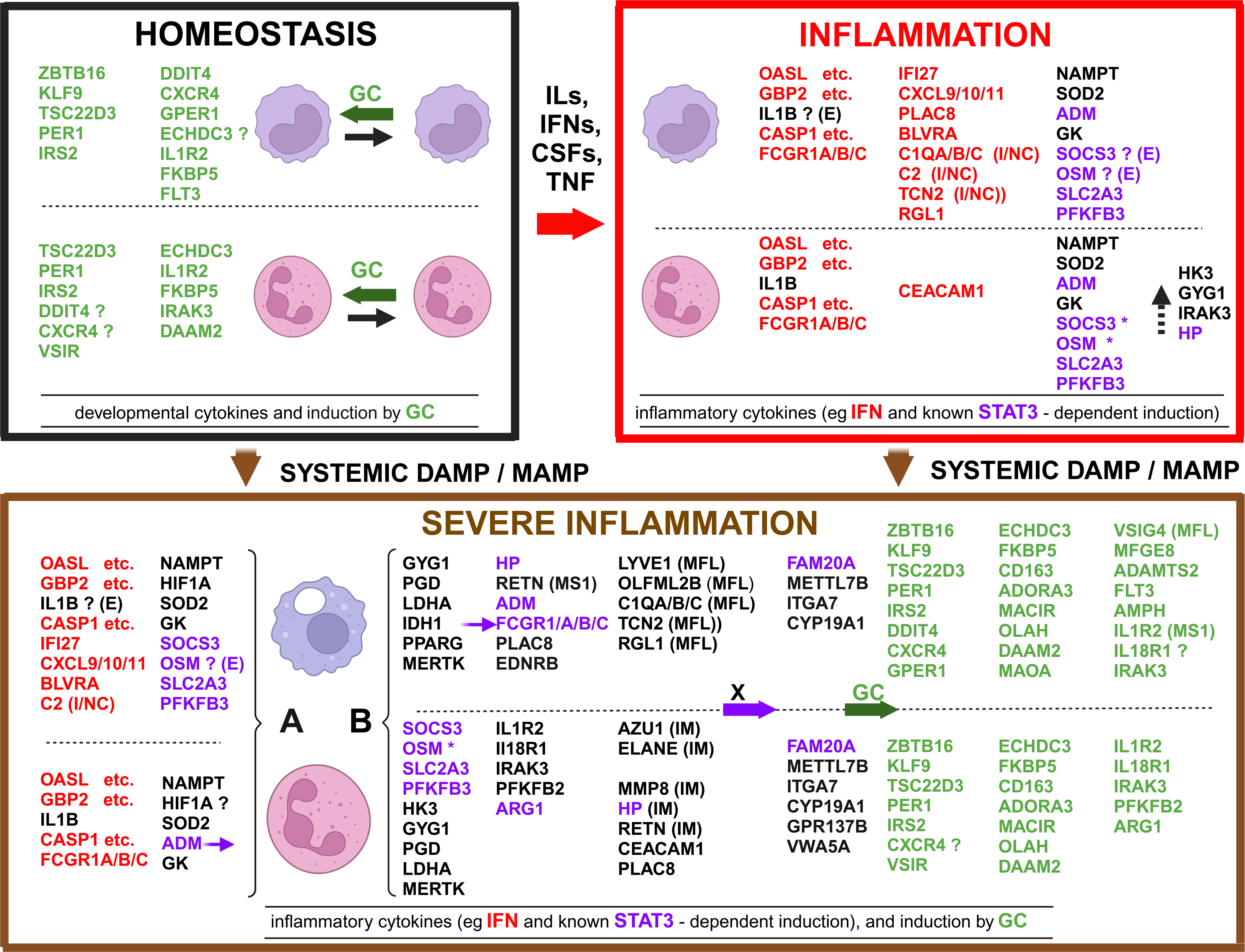
Comparing gene induction in myeloid cells, in homeostasis, inflammation, and severe inflammation. Gene induction in homeostasis, mild inflammation, and severe inflammation, in monocytes and neutrophils, shown for GC-induced genes (including biomarker sets GC-1 and GC-2), and for relevant marker genes induced during inflammation. The schematic combines results from correlation and differential expression analysis using different informative datasets (see main tekst). Circadian GC-induced gene expression was found during homeostasis in monocytes and neutrophils but with some differences, such as no detection of KLF9 and ZBTB16 in neutrophils, or IRAK3 in monocytes. Inflammation caused by mild respiratory viral infection, results in elevated blood cytokines, which induce gene expression in monocytes and neutrophils. Induction of marker gene IL1B is NF-kB-dependent, while induction of other genes involved interferon or known STAT3-dependent expression. Induction of IL1B, SOCS3, and OSM, as seen in separated monocytes and neutrophils was due to early gene activation by cell isolation procedure (marked: **? E**). In case of neutrophils, *in vivo* induction could be seen for these genes using whole blood datasets, instead (marked: **\***). Gene induction involving interferon was visible in intermediate / non-classical monocytes (**I/NC**), e.g. for C1QA/B/C genes. Less frequent upregulation, and upregulation much lower than in severe inflammation, occurred for a number of genes (eg GYG1) as indicated by a dashed vertical arrow. Severe inflammation in trauma or sepsis results in systemic release of DAMPs and MAMPs, and cytokines, which broadly induced 2 patterns of gene expression, one more similar to mild inflammation (**A**), the other more specific to severe inflammation (**B**). Macrophage-like monocytes (MFL) and immature neutrophils (IMM) were cell types characteristic for severe inflammation. Several genes were highly upregulated in severe inflammation, including FAM20A, due to a unknown inducer (combination) marked **X**. Elevated GC resulted in strong gene induction depending on cell type. Genes expressed in monocyte state MS1, and in macrophage-like monocytes (MFL) in sepsis as indicated.

Upregulation of C1QA/B/C genes and RGL1 in monocytes, and CEACAM1 in neutrophils, no longer involved induction by interferon (Figure S8, S10), as was also observed for whole blood data (Figure 6A). Broadly correlated expression of genes IL1B, GK, NAMPT, and SOD2 in monocytes and neutrophils, as seen in mild inflammation (Figure S7, S9), was also present in severe inflammation (Figure S8, S10). Changes in gene expression during sepsis, relative to healthy controls, were generally much larger in neutrophils than in monocytes, also in many cases of shared upregulation, as visible in dataset GSE60424 (Linsley et al., 2014).

Neutrophils reached higher relative expression levels of genes GYG1, HK3, PGD, and LDHA in glucose metabolism (Figure S10), than did monocytes, whereas monocytes reached higher relative expression levels of PPARG (Figure S8). Specific and strong upregulation in monocytes, only in severe inflammation, occurred for genes OLFML2B, EDNRB and LYVE1, the latter also constitutively expressed in neutrophils (Figure S8). A number of genes with absent / low basal expression in health and mild inflammation was strongly upregulated in both monocytes and neutrophils during severe inflammation, including gene FAM20A (Figure S8, S10). FAM20A is prominently present in an IL-10 driven gene signature *in vitro*, in monocytes ((GSE59184 (Montoya et al., 2014), GSE43700 (Teles et al., 2013), GSE47122 (Italiani et al., 2014), GSE146438 (Gorby et al., 2020); GEO2R, and supporting data (Figure S8)), M2c macrophages (Lurier et al., 2017), and IL-10 induced tolerogenic dendritic cells (GSE44719 (Banchereau et al., 2014), GSE45466 (Braun et al., 2013); GEO2R, (Avancini et al., 2023)). These experiments using IL-10 indicated STAT3-dependent induction of FAM20A *in vitro*, but the relevant inducer(s) *in vivo* remained unknown. Larger clusters of GC-regulated gene expression, beyond the genes shown earlier (Figure 3C, 3D, 3E, 3F), were evident from the data on separated monocytes and neutrophils (Table S7, S8, S9, S10). Genes MFGE8 in monocytes, and PFKFB2 in neutrophils, upregulated in severe inflammation, indicated a regulatory role for GCs in efferocytosis, and glucose metabolism, respectively.

### Frequent upregulation of set GC-2 genes early and late after severe injury

In case of severe injury, longitudinal data provide a precise time point for the start of severe inflammation. Such data might show whether start of gene induction by GC would correlate with the ordered appearance of different cell types. From the dataset on genomic storm occurring after critical injury (Xiao et al., 2011), neutrophil bands appeared in circulation immediately after injury, while later, immature neutrophils likely corresponded with myelocytes and metamyelocytes originating from emergency granulopoiesis (Figure 8, upper panels). Longitudinal changes in relative gene expression in neutrophils, and monocyte were visible, such as a decrease in expression of MME (CD10), a marker gene for old mature neutrophil, and of CD86, a marker gene for antigen presenting monocyte (Figure 8, row 2 panels). Gene marker PPARG for macrophage-like monocyte became upregulated compared to healthy control (Figure 8, row 2 panels). Induction by GC of genes in set GC-2 was frequently visible before, and after appearance of cells from emergency granulopoiesis, both in monocytes (eg VSIG4, ADAMTS2), and in neutrophils (eg OLAH, DAAM2). Although induction of GC-2 genes was frequently observed, a contribution of GC in treatment remained possible, since GC usage was not reported by Xiao et al (2011). VSIG4 expression peaked earlier than ADAMTS2, suggesting a different course for monocyte subtypes after injury (Figure 8, row 3 panels). Taking a similar approach as in case of selecting gene sets GC-1 and GC-2, several relevant gene sets for monitoring inflammation were selected using whole blood gene expression correlation and differential gene expression data, as detailed (Figure S11, and Figure S12). A gene set SIM (for Severe Inflammatory Myeloid cells, including GYG1, n = 16) was selected from a large group of highly upregulated genes in severe inflammation, especially in neutrophils, unrelated to GC induction (Figure S11). A related set FAM20A (n = 4) consisted of genes co-induced specifically in severe inflammation, also unrelated to GC induction (n = 4) (Figure S11). Gene sets were also used corresponding to induction by interferon gamma (n = 23, including GBP1), and interferon alpha (n = 8, including OASL), which were frequently upregulated in mild and in severe inflammation (Figure S12). Additionally, a gene set corresponding to T cells (n = 13, including LEF1, with bias for naive T cell) (Figure S13), was used to follow decreased percentages of T cells during inflammation. Upregulation of SIM set genes occurred immediately after injury, closely followed by upregulation of the FAM20A set, while relative expression of module T cells in blood was decreased instead. Relative expression of T cell module and SIM set was still different from healthy controls at week 4, indicating continued systemic severe inflammation in a number of patients. Combined expression of genes in set GC-2 was increased in most patient blood samples, expression of gene module interferon gamma was relatively unchanged, while expression of module interferon alpha was decreased (Figure 8, lower panels).

**Figure 8.**
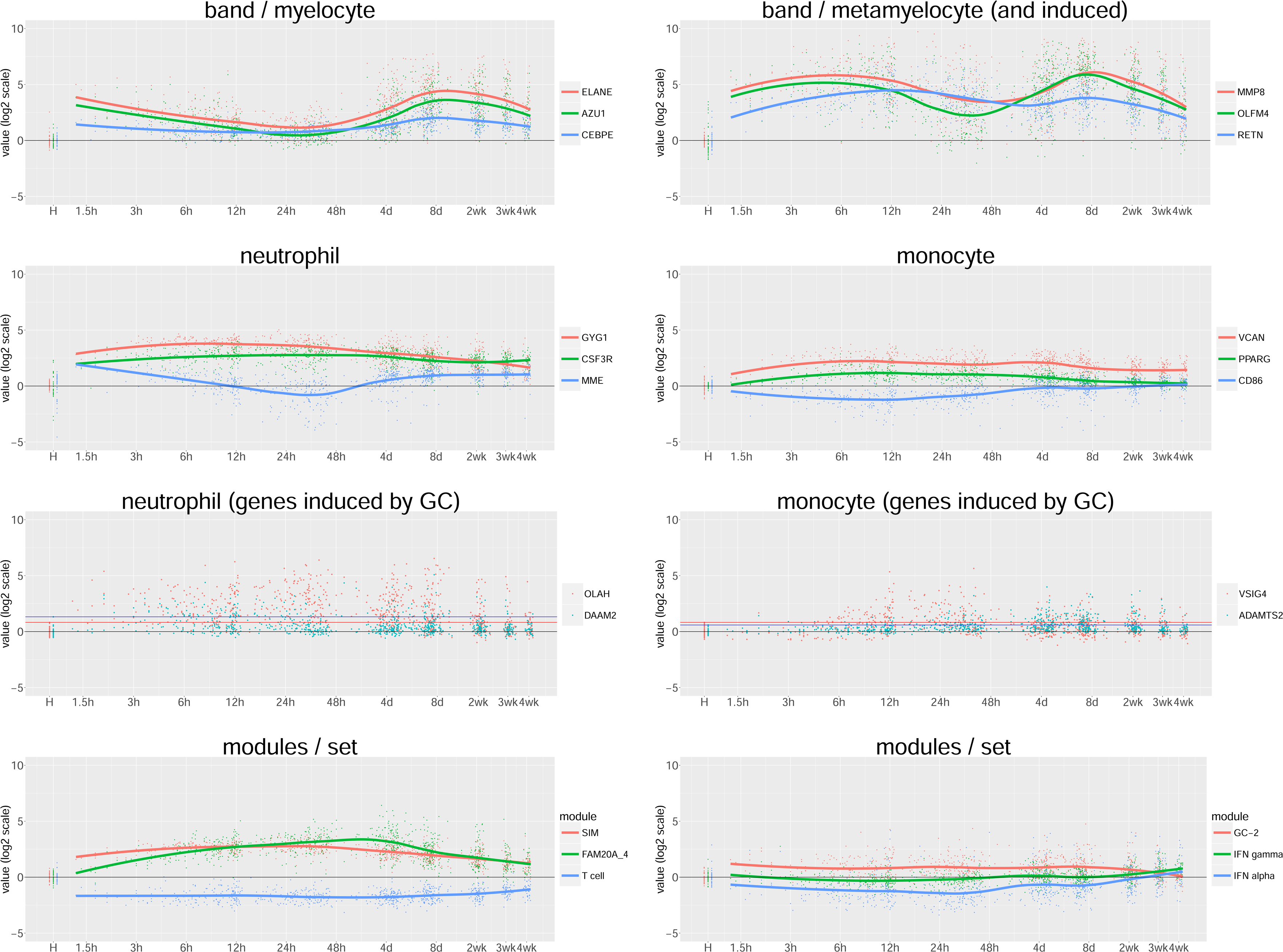
Longitudinal gene expression during severe inflammation after trauma. Expression of marker genes, GC inducible genes, and relevant gene sets in whole blood, after trauma, and compared to healthy controls (GSE36809 (Xiao et al., 2011)). Cell type specific expression and inductions of different genes as indicated in panels. Gene sets include SIM (severe inflammatory myeloid, n= 16), FAM20A_4 (n = 4), naive T cell (T cell, n= 13), GC- 2 (GC- inducible, n= 9), induction by interferon gamma (IFN gamma, n= 23), and induction by interferon alpha (IFN alpha, n = 8). Expression values and time after injury shown at log 2 scales (h, hours; d, days; w, weeks). Mean value for healthy controls (H) was set at zero. In case of GC- induced genes OLAH, DAAM2, VSIG4, and ADAMTS2, maximal expression values in healthy controls, which possibly still correspond to gene probe background levels, are indicated by horizontal lines. Expression values for gene sets were calculated using adequate gene probes, with subtraction of background / basal expression, and adjusting for different gene expression ranges as detailed in methods.

### Upregulation of GC-2 biomarker set expression by treatment with exogenous GC

Set GC-2 was tested as a transcriptomic biomarker using whole blood data with documented GC treatment. For a more complete description of systemic inflammation, gene sets T cell, SIM, FAM20A (n = 4), interferon gamma, and interferon alpha were used again as additional features. Adequate gene probes were selected for each gene in case of Affymetrix GPL570 and Illumina GPL10558 platforms, (Table S5), confirmed by expression correlation clustering using representative datasets (Figure S13). In case of set GC-1, gene PER1 was not included in gene set expression comparison, because of a single suboptimal probe present on the Illumina microarray platforms. Gene modules / sets were used as transcriptomic biomarkers using a measure of combined gene expression in the set (see methods). Expression value distributions were skewed and often with many outliers in case of the inducible modules (Figure 9). Increased expression of GC-1 and GC-2 sets was visible upon systemic GC treatment in burn- and septic-shock (Figure 9A, 9B). Increases in GC-1 and GC-2 expression in SLE and RA patients being treated with GCs (Figure 9C, 9D) agreed with results by Hu et al. (2018), who used an 8 gene molecular signature to monitor GC responses upon GC treatment. Upregulation of set GC-2 relative to GC-1 was much stronger in GC-treated burn shock and septic shock, than in SLE or RA. Expression of SIM and FAM20A gene sets was high in burn shock and septic shock, but relatively unchanged by GC treatment (Figure 9A, 9B). Interferon alpha and gamma modules were upregulated in SLE as expected (Figure 9C), while a marked decrease was visible in case of burn shock (Figure 9A), possibly related to more general shifts in gene expression in myeloid cells.

**Figure 9.**
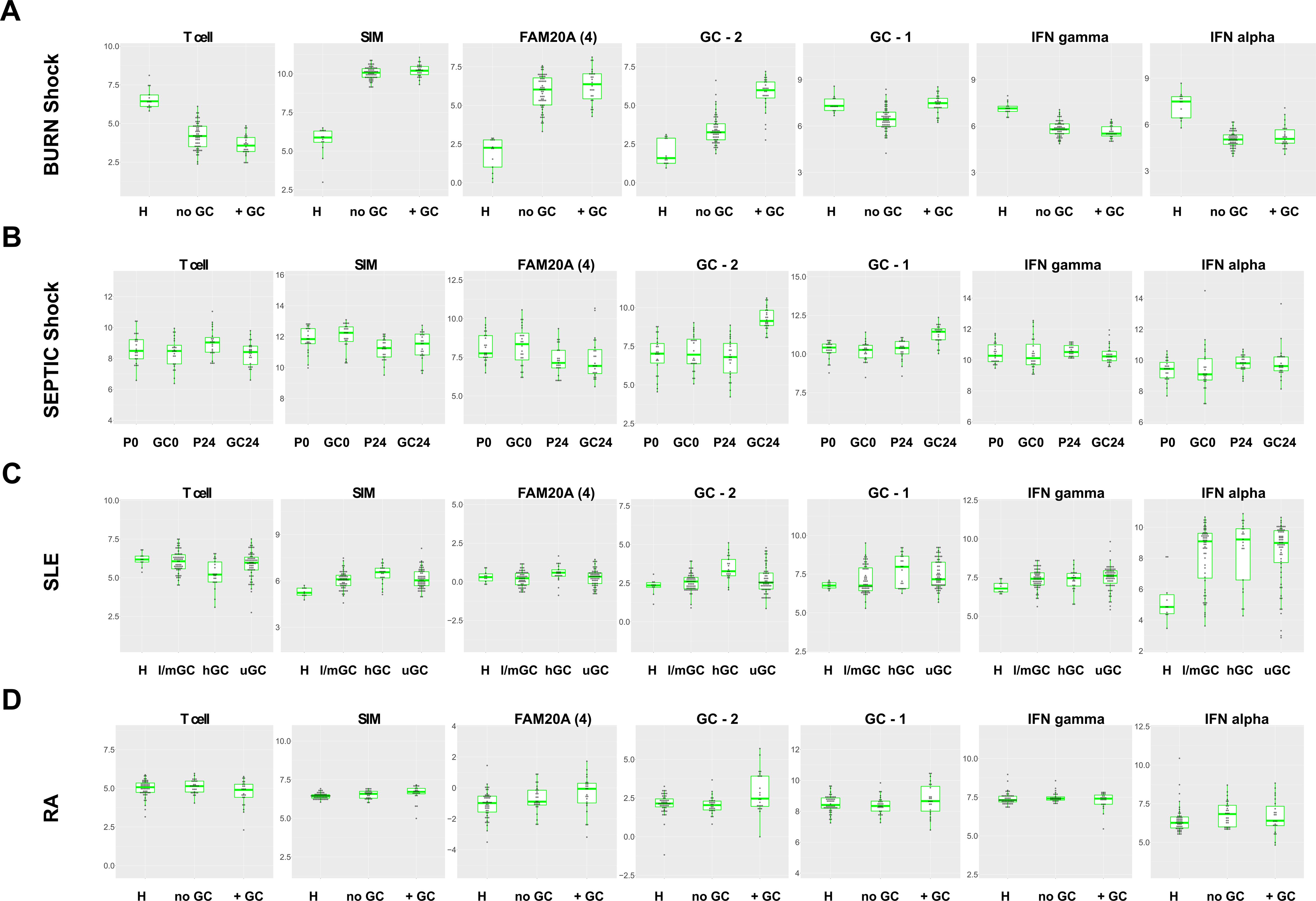
Distribution of gene sets expression in severe inflammation and autoimmune disease. Increase of GC-1 and GC-2 gene set expression upon GC treatment in patients with burn shock, septic shock, systemic lupus erythematosus (SLE), and rheumatoid arthritis (RA). Included for comparison are gene sets T cell, set severe inflammatory myeloid (SIM), set FAM20A, (4 genes including FAM20A), and sets interferon gamma (IFN gamma), and interferon alpha (IFN alpha). Datasets used different assay platforms, and log fold change between expression values in one dataset are not directly comparable to log fold changes in other datasets. Y-axis values on log-2 scale, with interval of 9 log 2 units for each panel. Shown are box plots with added binned dot plots. **A** GSE77791 (Plassais et al., 2017), healthy (H), severe burn shock (1-5 days, excluding 7 days samples (no GC), severe burn shock with hydrocortisone treatment (1-5 days, excluding 7 days samples (+ GC). **B** GSE106878 (Kolte, 2020), septic shock placebo 0h (P0), septic shock hydrocortisone 0h (GC0), septic shock placebo 24h (P24), septic shock hydrocortisone 24h (GC24). **C** GSE110174 (Hu et al., 2018), healthy (H), SLE low and medium prednisone usage (l/m GC), SLE high prednisone usage (hGC), SLE unknown prednisone usage (uGC). **D** GSE117769 published by Goldberg and co- workers (2018), healthy control (H), RA without GC treatment (no GC), RA with GC treatment (+ GC).

### Upregulation of GC-2 set by endogenous cortisol during severe inflammation in different diseases

Next, whole blood transcriptomic data covering many different diseases, and preferably including healthy controls, were examined by plotting gene set expression value distributions for patient groups. To allow a better comparison between datasets, the same adequate probes were used again in case of Affymetrix GPL570 and Illumina V3, and V4 (GPL10558) platforms (Table S5). Value distributions were plotted in the same range of 9 log2 units for all datasets, and datasets were grouped together by assay platform, including RNA-seq (Figures S14, S15, S16, S17, S18). The distributions for different sample groups were ordered within an experiment, and tentatively between experiments using the same platform, and between platforms, for use in an overview (Figure 10). Upregulation of SIM set genes was high in severe injury, sepsis, SIRS, severe COVID-19, Kawasaki disease, severe malaria, infectious diarrhea, lung cancer, active tuberculosis, IBD, and SJIA, very low in mild seasonal respiratory viral infection, and absent in latent tuberculosis, and asthma, with strength of differential expression and frequency of cases depending on disease (Figure 10, and Figures S14, S15, S16, S17, S18). Upregulation of FAM20A (n = 4) set genes generally accompanied upregulation of the SIM set genes, and was absent in mild respiratory viral infection, latent tuberculosis, and asthma. Expression of gene module IFN gamma was relatively high during active tuberculosis, active sarcoidosis, and leishmaniasis, while gene induction by IFN alpha was relatively high in acute respiratory viral infection, mild influenza, rotaviral diarrhea, and SLE, as expected (Figure 10). Upregulation of set GC-2 was relatively high in sepsis, and in severe COVID-19, besides GC-treated burn shock, and was also detected in additional diseases that are frequently treated by GC, namely severe asthma (exacerbation), JIA, SJIA, sarcoidosis, and IBD, besides SLE and RA (Figure 10, and Figures S14, S15, S16, S17, S18). Upregulation of set GC-2 was also seen in severe inflammation, in diseases that are not commonly treated with GCs, such as severe malaria, and infectious diarrhea, as well as in examples of diseases with explicitly documented absence of GC treatment, besides burn shock, namely in case of acute Kawasaki disease, and sepsis (Figure 10). Gene expression clustering of 19 GC-induced example genes, including set GC-2 was compared between healthy controls, severe malaria, infectious diarrhea, GC-untreated Kawasaki disease, and another example of GC-untreated septic shock (GSE110487, RNA-seq, (Barcella et al., 2018)), to check co-upregulation of the set. Relatively strong clustering of most GC-2 set genes, depending on disease, (Figure S19) indicated that GC-2 set genes were indeed co- regulated, as in case of GC-untreated septic shock and burn shock (Figure 4B, 4E, and 4F).

**Figure 10.**
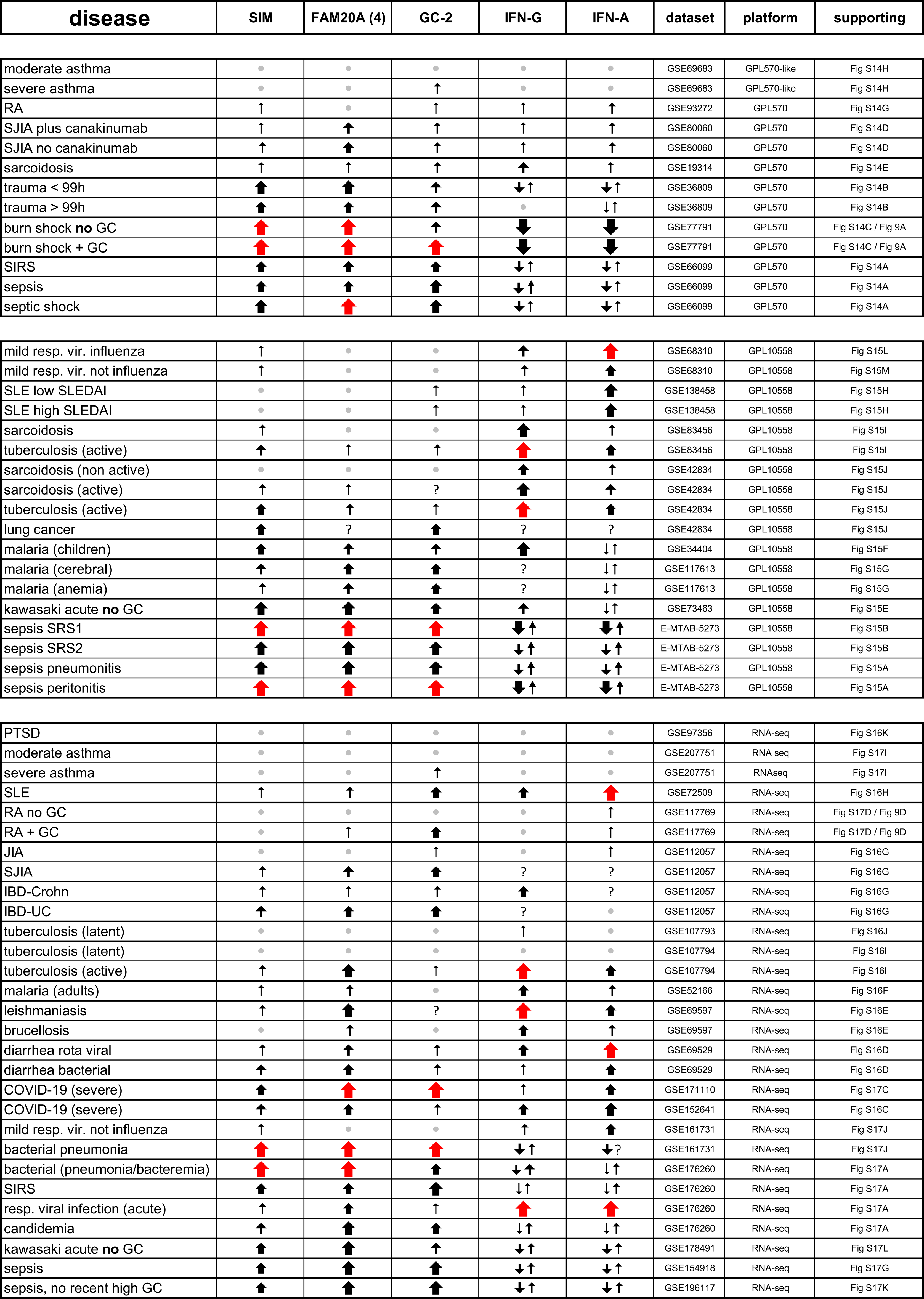
GC-2 gene expression is upregulated by endogenous and exogenous GC depending on disease. Overview of gene expression in whole blood during different diseases and upon treatment with GCs using 5 relevant features; a gene set for severe inflammatory myeloid (SIM), a gene set of 4 induced genes including FAM20A (FAM (4)), a gene set GC-2 for induction by GC during severe inflammation (GC-2), and gene sets for induction by either interferon gamma (IFN-G), or interferon alpha (IFN-A). Gene set expression values were plotted in ranges of 9 log 2 units and distributions compared within, and between experiments for platforms GPL570, GPL10558, and RNA sequencing data, and tentatively ordered according to deviation from healthy controls. Red arrows indicate maximal upregulation from healthy control for each gene set. Decreases in basal gene set expression, as frequently seen in case of interferon gamma and alpha induction in severe inflammation, may occur because of shifts in cell percentages, and do not necessarily indicate gene repression. Datasets, different platforms, and supporting figure panels as shown.

This analysis showed a transcriptomic signature of endogenous cortisol during severe inflammation for different diseases.

### Adjustment of GC biomarker sets for different diseases

Gene sets GC-1 and GC-2 partially overlapped with existing whole blood transcriptomic biomarkers for guiding GC replacement therapy in Addisons disease (Sævik et al., 2021), and pharmacodynamic monitoring of therapeutic GC in SLE, and RA (Hu et al., 2018; Northcott et al., 2021). To adjust biomarker gene sets for application in different diseases, we compared correlation characteristics, and fold upregulation for all reported genes separately, using whole blood RNA-seq datasets. These data encompassed the effect of circadian GC in health (GSE113883 (Braun et al., 2018)), GCs in sepsis (GSE154918 (Herwanto et al., 2021)), and in gastroenteritis (GSE69529 (DeBerg et al., 2018)), systemic GC usage in SLE (GSE72509 (Hung, Pratt, Sundararaman, Townsend, et al., 2015)), RA (GSE117769 by Goldberg and co- workers, 2018), severe asthma (GSE207751 by Ginebaugh and co-workers, 2022), JIA, SJIA, IBD (GSE112057 (Mo et al., 2018)), and severe COVID-19 (GSE171110 (Lévy et al., 2021), GSE157103 (Overmyer et al., 2021), GSE206264 (López-Martínez et al., 2023)), as well as the effect of high endogenous GC in GC-untreated severe COVID-19 (GSE168400 (Amado- Rodríguez et al., 2022), GSE197204 (López-Martínez et al., 2023)) (Table S6). Gene expression correlation profiles were obtained (Table S6), compared to each other (Figures S20, S21, S22, S23, S24), and checked for the presence of GC signature genes (Table S6, and Figure S25). Fold induction was determined by comparing with healthy controls, where available, and by examining gene expression value distribution range in disease (Table S6, and Figure S25). This analysis also highlighted inadequate annotation of gene ADORA3 on different assay platforms. Genes ADORA3 and TMIGD3 are chromosomal neighbors, and produce different (hybrid) transcripts and proteins (Ranjan et al., 2017), which are not always detected separately, or named correctly on different platforms. Platforms Affymetrix GPL570 and Illumina GPL10558 each contain 2 ADORA3 probesets and no TMIGD3 annotated probesets. Both platforms detect different transcripts according to Ensemble (Harrison et al., 2024), one corresponding with ADORA3, and expressed in eosinophils, another containing TMIGD3 sequence, and GC inducible. The specific probesets used in set GC-2 for both platforms corresponded to TMIGD3 sequence. Best marker genes for circadian GC action in health included set GC-1, additional genes such as FKBP5 and ECHDC3, and partially overlapped with “time telling” genes used in predicting circadian phase in health (Braun et al., 2018; Hughey, 2017; Laing et al., 2017), (Figure 11). Suitable marker genes for monitoring GC action in asthma, RA, and SLE were ZBTB16 (in set GC-1), set GC-2, and additional genes such as FKBP5, ALOX15B, IL1R2, and ARG1, and included most previously reported GC biomarker genes for SLE and RA (Figure 11). MAOA expression in reticulocytes represented an occasional confounder for GC induction. In general, the occurrence of severe inflammation in disease, and associated variability in neutrophils, limited the choice of biomarker genes, especially in severe COVID-19 and sepsis, e.g. excluding IL1R2 (Figure 11).

**Figure 11.**
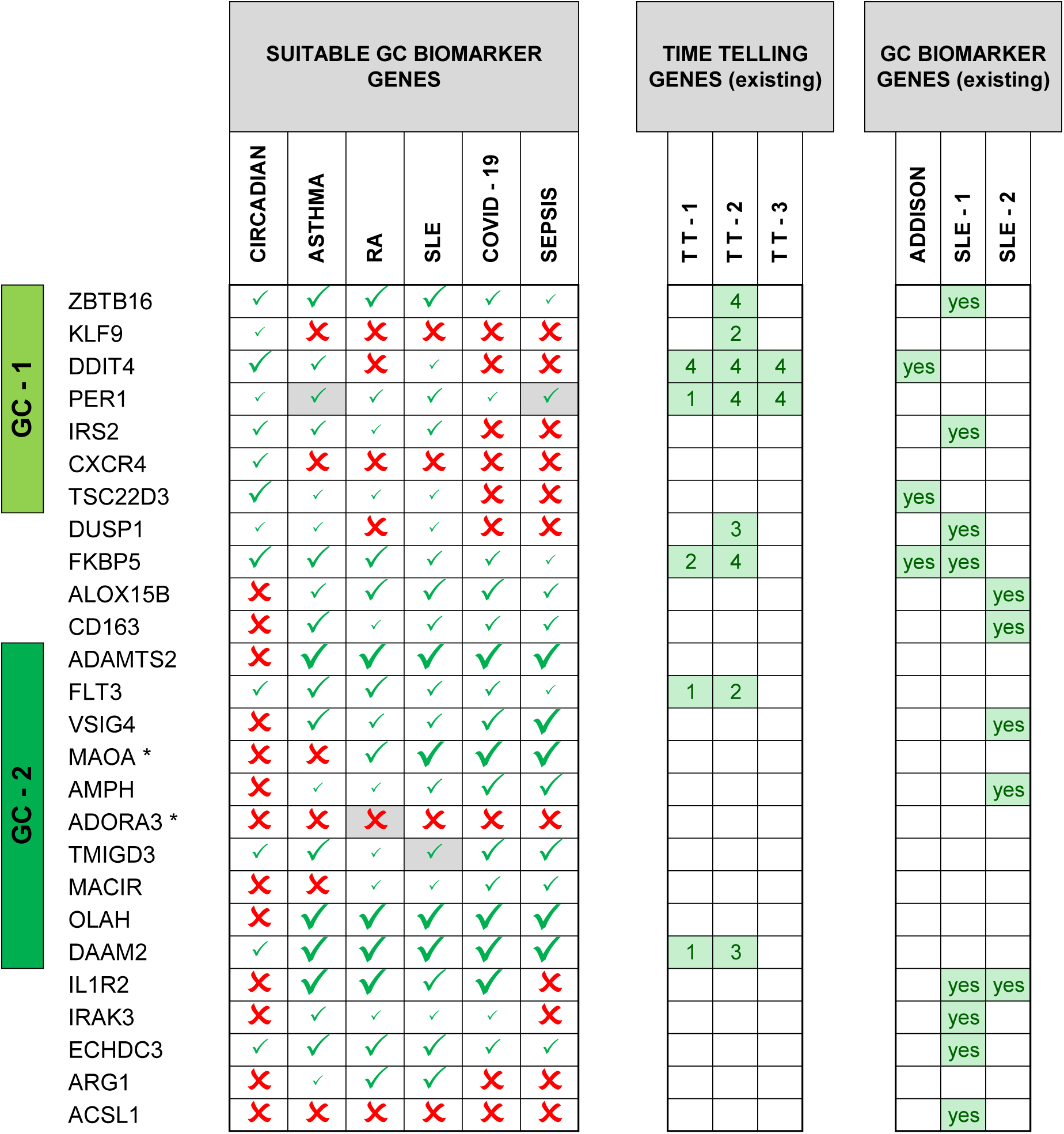
Evaluating biomarker genes of GC action in whole blood in different diseases. Left panel, scoring suitable biomarker genes of GC action in whole blood in health, and different diseases, based on induction strength and correlation analysis using relevant RNA- seq datasets (see main text). Included are all genes in set GC-1 and set GC-2, gene ARG1, and additional genes present in published GC biomarker gene sets. Grey marking, genes absent in single dataset, and taken from supplementary datasets; GSE69683 (asthma), GSE120178, and GSE129705 (both RA), GSE139940, and GSE110685 (both SLE), and GSE161731 (subset bacterial pneumonia and sepsis). Asterisk for ADORA3, and MAOA; see main text. Middle panel, genes present in whole blood transcriptomic sets of “time telling” genes identified by machine learning. In a 15 genes set for monitoring the circadian clock in blood (Hughey, 2017), (TT-1), in 234 genes identified as phase markers by Laing et al. (2017), (TT-2), and in a a set of 41 predictive genes for circadian state (Braun et al., 2018), (TT-3). Relative strength indicated from 1 to 4, based on coefficients in 2 principal components used for prediction (TT-1), number of times identified as circadian phase marker using 4 different methods (TT- 2), and high frequency of selection in predictor in different training samples (TT-3). Right panel, genes present in whole blood transcriptomic biomarker sets for guiding GC replacement therapy in Addison’s disease (Sævik et al., 2021), and monitoring response to GC treatment in SLE and RA (Hu et al., 2018), (SLE-1), and SLE (Northcott et al., 2021), (SLE-2).

## DISCUSSION

### Aim of study and short summary of results

In this study, we aimed to develop gene expression biomarkers for GC action on blood cells. We obtained gene signatures of GC induction by using correlated expression with several well-known GC target genes, used as queries, in meta-analysis of collections of public blood transcriptomic datasets. Different signatures were found depending on the absence or presence of severe inflammation, as in case of sepsis and critical injury. These 2 signatures, which both showed good overlap with GC upregulated genes in immune cells *in vitro*, originated from *in vivo* expression, thereby providing an alternative starting point for selecting biomarker genes. GC signatures 1, with different cell type biases, originated from circadian gene induction by GC in lymphocytes, neutrophils, and monocytes. From this signature, we selected a set of genes (GC-1) as best biomarkers for GC action, for use on whole blood in the absence of severe inflammation. Set GC-1 could be used efficiently, to detect induction by endogenous circadian cortisol, and expression was not upregulated during mild inflammation. GC signatures 2, also with different cell type biases, originated from lymphocytes, and from alternatively activated monocytes and neutrophils in severe inflammation. Upregulation of key signature genes expressed in myeloid cells was visible in whole blood, upon GC treatment in burn shock, and septic shock. We demonstrate that expression of a selected set of GC signature 2 genes (GC-2) was well suited to detect gene induction in blood upon systemic GC treatment in SLE, RA, asthma, in septic shock, and in burn shock. GC signature 2 was found in many more different inflammatory illnesses, including Kawasaki disease, severe malaria, severe COVID-19, and gastroenteritis, resulting from both exogenous and endogenous GC. Use of set GC-2 indicated different frequencies and strength of induction in patients depending on illness. Using meta-analysis, we also provide an overview of gene expression, in homeostasis, inflammation, and severe inflammation in circulating monocytes and neutrophils, more generally.

### Meta-analysis of gene expression correlation is a useful method

Differential gene expression in blood upon *in vivo* GC or ACTH treatment has been widely studied in humans and animals. The change in blood cell percentages caused by these treatments enormously complicates the interpretation of the transcriptomic data. Potential GC- regulated DEGs in these experiments are usually identified from existing literature, or by comparing *in vivo* with *in vitro* gene induction results, for example in PBMCs (Hu et al., 2018). More exceptionally, a set of co-expressed GC-induced genes in blood of ACTH injected pigs was directly identified by cluster analysis using a single experiment (Sautron et al., 2015). We noticed induction signals originating from circadian and stress-related changes in endogenous cortisol, and sometimes also from GC usage, leading to gene expression correlation profiles in many blood transcriptomic datasets, which provided a useful starting point for finding candidate GC-regulated genes. It indicated that a search would not necessarily be limited to GC administration experiments or studies with documented GC usage, and that Pearson’s correlation might be used in meta-analysis to obtain robust gene expression correlation profiles. In case of the initial GC signatures 1 and 2, datasets were selected from a total number of 375+ blood transcriptomic datasets, based on the quality of correlation hits. Subsequent gene expression correlation profiles on whole blood were obtained in meta-analysis using transcriptomic datasets collections based on experimental description, to ensure that GC-driven gene expression was separately observed for circadian GC, and GC in severe inflammation. The recovery of known GR target genes in GC signatures, many recurrent candidate genes, and the large overlap with GC upregulated DEGs in blood cells in different *in vitro* and *in vivo* experiments showed that the approach was generally very effective. Likewise, a large overall signature of GC-driven gene expression in blood and tissues, corresponded well with GC upregulated DEGs in immune cells and cell cultures observed *in vitro*. A general advantage of using correlation is its independence from gene expression range and basal expression levels, or background signals often present in microarray data, allowing easier detection of GC-regulated genes at low-fold induction. A difficulty with correlation analysis was that GC-regulated DEGs may drop down in gene expression correlation profiles due to overriding alternative correlations, and thus escape detection.

### Circadian GC-driven gene expression

Circadian gene expression in whole blood has been well documented in experiments on insufficient sleep and mistimed sleep (Archer et al., 2014; Arnardottir et al., 2014; Möller- Levet et al., 2013). Circadian rhythms occurring in differential (relative) blood counts (Ackermann et al., 2012; Lange et al., 2022) can be expected to result in mainly cell type- specific circadian gene expression in whole blood. Neutrophil and NK numbers go up at day time, monocyte numbers remain relatively unchanged, while T and B cell numbers go down at early day time, regulated by cortisol via the HPA axis, and catecholamines via the sympathetic nervous system (Lange et al., 2022). It is likely that higher percentages of NK cells and neutrophils, coinciding with higher gene induction by cortisol, lead to the corresponding cell type biases in blood-based GC signatures 1. Using dataset collections of separated monocytes, neutrophils, and PBMCs allowed detection of circadian GC-driven gene expression in different cell types more precisely. Several methods exist to predict the circadian phase from transcriptomic measurements of whole blood (Braun et al., 2018; Hughey, 2017; Laing et al., 2017) and monocytes (Wittenbrink et al., 2018). A glucocorticoid driven network links many of the top ranked biomarker genes for circadian phase in whole blood, indicating gene induction by circadian GC either via local circadian clocks or directly (Laing et al., 2017). In agreement, among the circadian expressed, best “time telling” genes in these studies were many genes present here in GC signatures 1 and 2 (9/15 (Hughey, 2017); e.g. 13/76 (Laing et al., 2017); 3/40 (Braun et al., 2018); e.g. 10/34 (Wittenbrink et al., 2018)).

### GC-driven gene expression in monocytes and neutrophils

It has been argued that a better understanding of cell type and tissue specific effects of GCs, would be important for adjusting or developing new therapies (Quatrini & Ugolini, 2021) . Highly cell type specific gene induction upon *in vitro* GC treatment was reported in isolated immune cells and in proliferating non-immune cell cultures, and the global effects of GC in different cells were compared at the molecular pathway level (Franco et al., 2019). Here, we traced *in vivo* effects of endogenous GC on the transcriptome of lymphocytes, monocytes, neutrophils, muscle, skin, and fat, by using expression correlation with GC-induced query genes in meta-analysis. The focus here was on monocytes and neutrophils, since they contributed most to the GC-driven signatures detected in whole blood. Generally stronger signatures present in severe inflammation compared to health, might reflect higher GC levels and induction in severe inflammation than during circadian rhythm. Circadian GC-driven gene expression in health, which would be laborious to document using alternative approaches, showed differences between monocytes and neutrophils, and mostly represented a subset of genes induced in severe inflammation. Finding VSIR, encoding VISTA, as a novel GC-regulated gene in neutrophils added to the importance given to VISTA as a possible therapeutic target in managing cytokine storms (ElTanbouly et al., 2021), Similarly, finding GPER1, encoding the GPCR estrogen receptor, as induced in monocytes, pointed to the anti- inflammatory activity of estrogen on monocytes (Pelekanou et al., 2016), being modulated by cortisol. Gene expression correlation profiles for monocytes in severe inflammation shared many genes, reported as GR-targeted DEGs in monocytes and macrophages *in vitro* (Wang et al., 2019), or as GC-regulated DEGs *in vivo*, in isolated monocytes from giant cell arteritis patients (Estupiñán-Moreno et al., 2022). GC treatment of isolated monocytes triggers an extensive transcriptional program, directing monocyte to macrophage differentiation to an alternatively activated state with anti-inflammatory phenotype (Ehrchen et al., 2007; Ehrchen et al., 2019; Heideveld et al., 2018; Wang et al., 2019). GC treatment of monocyte-derived macrophages also induces alternatively activated macrophage subtypes with roles in wound repair, tissue remodeling and efferocytosis (Abdelaziz et al., 2020; Desgeorges et al., 2019; Ehrchen et al., 2019; Roszer, 2015). Important genes functional in tissue repair (ADAMTS2), (Ehrchen et al., 2019), and efferocytosis (C1QA/B/C, MFGE8), (Ehrchen et al., 2019) were present in gene expression correlation profiles from monocyte separations in severe inflammation. Notable with respect to wound healing was regulation by GC of genes ANG, RNASE1, and RNASE4, encoding angiogenic factors. In this way, the response of blood mononuclear phagocytes to GC in severe inflammation resembled alternative activation. Due to tight gene expression correlation clustering caused by induction by GC, assigning gene expression to specific mononuclear phagocytic cell types, such as MS1 monocytes, non- classical monocytes, macrophage-like monocytes, and/or myeloid dendritic cells remained difficult, and would require precise analysis of single cell RNA sequencing data on sepsis.

Gene expression correlation profiles for neutrophils in severe inflammation also shared many genes, reported as upregulated DEGs in neutrophils treated by GC (Franco et al., 2019). The GC signatures from blood and isolated neutrophils indicated important effects of GCs on inflammatory neutrophils during severe inflammation, for example modulation of IL1 and IL18 signaling (e.g. by upregulation of IL1 decoy receptor IL1R2, IL18R1, and IRAK3), modulation of leukotriene chemotaxis and synthesis (upregulation of LTB4R encoding leukotriene B4 receptor, and upregulation of ALOX5AP/FLAP), and prostaglandin breakdown (upregulation of HPGD). Secretion of arginase 1, encoded by ARG1, has been implicated in immunosuppression by low density neutrophils in severe inflammation, and arginase derived from tumor infiltrating myeloid cells is an important factor in suppression of the cancer immune response (Grzywa et al., 2020; Mortaz et al., 2018). As ARG1 is also upregulated in neutrophils by *in vivo* dexamethasone treatment of severe COVID-19 (Sinha et al., 2022), and ARG1 is a GC receptor targeted gene in liver (Okun et al., 2015), our results strongly suggest it might be a GC target gene in neutrophils as well.

### Cell biology of monocytes and neutrophils in mild and severe inflammation

A very large quantitative and qualitative difference existed between blood gene expression during inflammation in mild respiratory viral infection, and in severe inflammation. Blood gene expression, specific to severe inflammation, is likely ultimately due to gene induction by systemic DAMPs and MAMPs, as in trauma and sepsis (Raymond et al., 2017; Relja & Land, 2020). Blood transcriptomics, including gene expression in monocytes and neutrophils, has been used to correlate gene expression with degrees of severity in COVID-19 (Wang et al., 2022). Immature neutrophils originating from emergency myelopoiesis are a heightened characteristic in severe COVID-19 and sepsis (Aschenbrenner et al., 2021; Kwok et al., 2023; Schulte-Schrepping et al., 2020). Based on their complete absence, we suggest that data on experimental, and prospective, seasonal respiratory viral infection, may represent a clean example of systemic mild inflammation, without a transition to severe inflammation as frequently occurring in COVID-19 patients. Mild inflammation as seen in uncomplicated respiratory viral infection was especially marked by inductions involving interferon, and also by upregulation of NF-kB-dependent IL1B, and STAT3-dependent SOCS3, as expected.

Upregulation of GK, encoding glycerol kinase, expressed in correlation with ACSL1 and AQP9, mainly in neutrophils in mild inflammation, might indicate increased triacylglycerol synthesis in support of lipid body biogenesis, which is important in the production of inflammatory mediators from arachidonic acid (Melo & Weller, 2016; Weller, 2016).

Upregulated expression of CDKN1C and C1QA/B/C genes was consistent with increased frequencies of intermediate monocytes present in blood in mild respiratory viral infection (Vangeti et al., 2023). Differential expression correlation of highly upregulated genes between inflammation in mild respiratory viral infection and in sepsis (as observed for CEACAM8 in neutrophils, and genes FCGR1A/B/C, and RGL1 in monocytes) indicated that genes may become targets for alternative induction pathways in sepsis, and hypothetically, switching from interferon to relatively strong induction by IL6.

Using single-cell RNA sequencing, 4 different monocyte states have been defined in sepsis including a population similar to non-classical monocytes named MS3, expressing high FCGR3A (CD16), and a CD14 +, HLA-DR low subpopulation named MS1. MS1 expresses RETN, and IL1R2, is expanded in sepsis and COVID-19, and originates from altered myelopoiesis in bone marrow (Reyes et al., 2020; Reyes et al., 2021). Additional marker genes can be found for MS1, and for MS3 (e.g. CDKN1C, C1QA/B/C genes) in the study by Reyes et al. (2020). Here, the existence of different gene expression clusters in monocytes in severe inflammation, with either C1QA/B/C genes or CDKN1C (non-classical-like), suggested important heterogeneity in MS3 cells. Of relevance, a macrophage-like cell type, separate from non-classical monocytes, has been characterized by single cell RNA sequencing of cerebrospinal fluid and PBMCs in tuberculous meningitis, expressing C1QA/B/C genes, SLCO2B1, LYVE1, as well as VSIG4, MSR1, and OLFML2B (Mo et al., 2024). Here, we show upregulated co-expression of these genes in monocyte separations, including VSIG4 and C1QA/B/C genes, and upregulation of macrophage marker genes LYVE1, and PPARG, indicating similar circulating macrophage-like cells, more generally in severe inflammation.

Juss et al. (2016) have described a large number of highly upregulated genes in neutrophil separations in ARDS compared to healthy controls. We found that many of these genes were not at all upregulated during inflammation in mild respiratory viral infection. Other genes such as GYG1 were upregulated in mild inflammation, also, but at lower fold change and lower frequency. This qualitative transcriptomic difference indicated the importance of distinguishing between mild and severe inflammation. Low density neutrophils (LDNs) containing immature and mature granulocytes expand in sepsis and COVID-19 (Mortaz et al., 2018; Uhel et al., 2017), (Kwok et al., 2023; Reusch et al., 2021; Schrijver et al., 2019; Schulte-Schrepping et al., 2020). Immature granulocytes were indicated here also, by strong correlated upregulation of proliferation and azurophilic granule marker genes. Neutrophils and monocytes in severe inflammation shared upregulation of many genes, including genes co-expressed with FAM20A, and many GC-regulated genes. Upregulation of CD163 and MERTK seen in gene expression datasets in severe inflammation agreed with flow cytometric data of both monocytes and neutrophils in sepsis (Groselj-Grenc et al., 2008; Guignant et al., 2013), and also indicated similar functional changes occurring in neutrophils and monocytes.

### Severe inflammation occurring in different illnesses

Upregulated DEGs in myeloid cells in severe inflammation were analyzed by gene expression correlation analysis, to derive more specific features. A SIM (Severe Inflammatory Myeloid) gene set (n = 16) was selected from a large group of upregulated genes in severe inflammation in myeloid cells, and probably remained more heterogeneous with respect to gene regulation. This gene set was well suited to detect severe inflammation in patient samples for many different diseases. Within the large group of upregulated genes, a small subcluster of genes co-expressed with FAM20A in whole blood was used as a potentially informative biomarker gene set (n= 4). Besides FAM20A, genes METTL7B, and CYP19A1 in the set are also upregulated in IL-10 induced M2c macrophages (Lurier et al., 2017). METTL7B and ITGA7 are neighboring genes on chromosome 12, which might suggest shared regulation of gene expression. It would therefore be interesting to determine if FAM20A (geneset) might qualify as a whole blood IL-10 induction biomarker *in vivo*. Another group of upregulated genes in severe inflammation in myeloid cells were GC inducible genes, from which gene set GC-2 (MACIR/C5orf30, ADORA3, DAAM2, OLAH, ADAMTS2, FLT3, VSIG4, AMPH, MAOA) was selected. GC treatment in septic shock and burn shock induced many genes in GC signatures 1, and 2 above the expression level reached by endogenous GC. Previously, upregulated DEGs in GC treatment in septic shock were not discussed as potential GC target genes, possibly since IPA (Ingenuity Pathway Analysis) at the time showed down-regulation of glucocorticoid signaling pathways instead (Wong et al., 2014). A small number of well- known GC target genes were reported as upregulated DEGs in GC treatment in burn shock (Plassais et al., 2017), but not identified within a larger cluster of co-expressed GC-regulated genes. Furthermore, upregulation of set GC-2 was frequent in GC-untreated Kawasaki disease, severe malaria, severe COVID-19, and infectious diarrhea, which indicated induction by endogenous GC, and not just by exogenous GC in treatment. These results were in good agreement, with increased serum cortisol present in critical illness more generally, and also in Kawasaki disease (Aso & Satoh, 2021; Sano et al., 2010), severe malaria (Enwonwu et al., 1999; Vandermosten et al., 2023), COVID-19 (Tan et al., 2020), and gastroenteritis (Rezai et al., 2022). Endogenous GC has been discussed as a possible factor in immunosuppression in sepsis (He et al., 2020; Vandewalle & Libert, 2020), although not commonly so (Fu et al., 2023; Hibbert et al., 2018; Liu et al., 2022; Ono et al., 2018; Padovani & Yin, 2024; Torres et al., 2022; Venet & Monneret, 2018). On the other hand, myeloid cells have been widely implicated in immunosuppression in severe inflammation, in case of monocytes (Avendaño- Ortiz et al., 2018; Mengos et al., 2019; Shalova et al., 2015), neutrophils (Darcy et al., 2014; Demaret et al., 2015; Hesselink et al., 2019; Mortaz et al., 2018; Reusch et al., 2021; Silvestre-Roig et al., 2019; Sinha et al., 2022), or both cell types (Reyes et al., 2021; Schrijver et al., 2019; Schulte-Schrepping et al., 2020; Uhel et al., 2017; Venet et al., 2021). Low expression of HLA-DR is typical for monocytes in sepsis, and is an important biomarker for immunosuppression (Cajander et al., 2013; Hibbert et al., 2018; Monneret & Venet, 2014; Torres et al., 2022). Low monocyte HLA-DR expression in septic shock, and after surgery was found to correlate with high serum cortisol in earlier reports, while treatment by GC *in vitro* downregulated HLA-DR (Kim et al., 2010; Tulzo et al., 2004). Here, we find a wide functional range of genes induced by GCs in circulating monocytes and neutrophils in severe inflammation. This likely pointed to an important role for endogenous GCs in immunosuppression in sepsis, via myeloid cells, which would be worthwhile to explore.

### Transcriptional biomarkers

Blood gene transcriptional modules of co-expression can be used as a basis for biomarker discovery (Chaussabel & Baldwin, 2014; Chaussabel et al., 2008). Here, we selected 2 gene sets GC-1, and GC-2 for use as blood transcriptomic biomarkers for GC action on cells, based on expression correlation and differential expression analysis of many public datasets.

Different sets of GC-induced genes have recently been selected for monitoring the response of blood cells to GC treatment in SLE, RA and Addison’s disease (Hu et al., 2018; Northcott et al., 2021; Sævik et al., 2021). Set GC-1 might represent a set suitable for use on whole blood in the absence of severe inflammation, similar to a set of 3 genes (TSC22D3, DDIT4, and FKBP5) proposed for guiding GC replacement therapy in Addison’s disease (Sævik et al., 2021). Set GC-1 genes are among the high ranking DEGs in PBMCs in GC therapy of Addison’s disease reported in a separate study (Chantzichristos et al., 2021), supporting their potential use as biomarkers. In case of inflammatory illnesses, GC-1 set genes would become less suitable for straightforward use on whole blood or PBMCs due to changes in cell percentages, and large shifts in differential gene expression in specific cell types, especially neutrophils, which tend to obscure the existing induction signal. Hu and coworkers developed a set of 8 biomarker genes for GC activity (FKBP5, ECHDC3, IL1R2, ZBTB16, IRS2, IRAK3, ACSL1, DUSP1), based on differential expression seen upon GC treatment both in health and in SLE, which can be applied to whole blood using qPCR (Hu et al., 2018). An alternative set of 5 genes (VSIG4, ALOX15B, CD163, AMPH, IL1R2), based on expression correlation with GC usage in both PBMC and whole blood datasets on SLE, was also tested for biomarker potential (Northcott et al., 2021). These selections apparently originated from induction by GC, in neutrophils and monocytes, respectively. Here, set GC-2 combined several genes highly induced by GC in monocytes and neutrophils in severe inflammation, and was adequate for detecting effects of GC usage in both autoimmune diseases, and in sepsis and burn shock. If required, the present data analysis would also allow a precise selection of GC biomarker genes expressed in either monocytes or predominantly in neutrophils, as well as extension of set GC-2 depending on disease. The more limited choice of suitable genes in sepsis compared to SLE, RA, and asthma might be a consequence of strong additional inductions, besides by GC, occurring in sepsis, e.g. in case of genes IL1R2, IRAK3, and ARG1.

Specific blood transcriptional endotypes in septic shock are associated with increased patient mortality in GC treatment, suggesting it would be important to account for heterogeneity using transcriptomics (Antcliffe et al., 2019; Wong et al., 2015; Wong et al., 2021). Serum IFN gamma / IL-10 ratio has also been proposed as a biomarker to help decide on GC treatment in septic shock (König et al., 2021). As impaired corticosteroid metabolism in sepsis is thought to be a factor in the outcomes of GC treatment (Annane et al., 2019), GC inducible genes TSC22D3 (encoding GILZ), and DUSP1 (encoding MKP1) are included with other biomarkers for stratification in clinical trial RECORDS on corticosteroid (GC + MC) sensitivity in sepsis (Fleuriet et al., 2023). In case of using whole blood transcriptomic data, set GC-2 would represent a more robust alternative biomarker, also in retrospective analysis, and for determining the transcriptomic effects of combined hydrocortisone (GC) and fludrocortisone (MC) treatment. In general, it would be worthwhile to relate set GC-2 expression to other measurements and clinical data, such as possibly immunosuppression by endogenous GC in sepsis, as well as by exogenous GC in treatment.

Relevant gene module / biomarker combinations have been used for patient stratification in subclasses, for example in SLE, bacterial infection, severe RSV infection, and sepsis (Altman et al., 2021; Banchereau et al., 2016; Banchereau et al., 2012; Northcott et al., 2021; Rinchai et al., 2020). In contrast, gene modules have not been applied in most other studies on stratification of sepsis in endotypes as reviewed (Komorowski et al., 2022; Pelaia et al., 2023; Tsakiroglou et al., 2023). A smart combination of biomarker features, closely corresponding to gene transcriptional modules, would allow a basic description of patient immunological status in different inflammatory illnesses. Gene induction by GC will likely be a relevant feature, as well as, for example, inductions by interferon alpha and gamma in a larger biomarker panel. In future, such panels might then be used for initial classification, and for immune status monitoring, building on more easily understandable features, than in case of existing sepsis endotypes.

## LIMITATIONS

GC-driven gene expression correlation profiles were determined here by the transcriptomic datasets present in a collection, and the GC-induced query genes used for obtaining a combined profile, both of which are choices, that can be made differently. Gene expression correlation offers an integral view of cellular processes, which are often not known to be connected. We were most interested in expression correlations with query genes directly caused by gene induction by GC, which necessitated a great deal of cross-checking of different gene expression profiles and comparison with *in vitro* and *in vivo* GC upregulated genes. In case of DEGs a common rule for cut-off is 2 fold change and adjusted p-value below 0.05, whereas selecting best candidate genes for GC induction from GC signatures remained difficult.

Changing percentages of cell types in blood hugely complicates blood transcriptomic analysis, as noted by many authors. Here, we developed biomarker gene sets for use on whole blood, since whole blood is easier obtained than separated cells in clinical practice, and because many datasets on whole blood are already deposited in public databases and available for analysis. Expression levels of these biomarker sets, especially set GC-1 for circadian GC action likely remain sensitive to changes in cell percentages, limiting their usefulness. It would be more accurate, and also easier, to develop GC transcriptomic biomarkers for use on data of separated cell (sub) types instead.

Clearly, there is a limit to novel insights that can be gained from analysis of bulk transcriptomics as performed here, regarding monocyte and neutrophil cell biology in homeostasis and during inflammation. Analysis of public single cell sequencing data would have allowed mapping (abundant) expression of GC-induced genes and marker genes to specific cell subtypes more precisely. It might also reveal whether different inductions can co- exist in the same cell, and infer trajectories. The main intention here was to place inductions by GC in a broader framework of gene expression in homeostasis, mild, and severe inflammation.

## Data Availability

All data produced in the present study are available upon reasonable request to the authors

https://www.umcutrecht.nl

https://www.ncbi.nlm.nih.gov/gds

https://www.ebi.ac.uk/biostudies/arrayexpress

https://github.com/ArthurJohannes/GCblood_repo

