## Supplementary material for "Glucocorticoid-driven gene expression in circulating monocytes and neutrophils in health and severe inflammation": Figure S6

RANK CORRELATION GC | EXPRESSION RATIO

In each table below, the first four value columns (MoH, MoS, NeH, NeS) are RANK CORRELATION GC; the remaining eight columns (MoH, MoS, NeH, NeS, LyH, LyS, MyH, MyS) are EXPRESSION RATIO.

### regulation of gene expression

| | MoH | MoS | NeH | NeS | MoH | MoS | NeH | NeS | LyH | LyS | MyH | MyS |
|---|---|---|---|---|---|---|---|---|---|---|---|---|
| TAF8 | 11 | 254 | 45 | 20 | 0,28 | 0,28 | 0,18 | 0,26 | 0,32 | 0,32 | 0,16 | 0,19 |
| KLF9 | 28 | 20 | 109 | 7 | 0,34 | 0,52 | 0,01 | 0,12 | 0,33 | 0,45 | 0,08 | 0,14 |
| ZBTB16 | 1 | 202 | 9381 | 12 | 0,39 | 0,46 | 0,05 | 0,11 | 0,41 | 0,4 | 0,08 | 0,1 |
| JDP2 | 29 | 43 | 428 | 182 | 0,26 | 0,27 | 0,28 | 0,19 | 0,02 | 0,03 | 0,51 | 0,44 |
| SAP30 | 17 | 11 | 51 | 17 | 0,36 | 0,53 | 0,05 | 0,06 | 0,12 | 0,18 | 0,28 | 0,42 |
| PHC2 | 32 | 15 | 1 | 76 | 0,06 | 0,15 | 0,4 | 0,38 | 0,05 | 0,06 | 0,42 | 0,48 |
| PER1 | 22 | 1309 | 53 | 88 | NA | NA | NA | NA | NA | NA | NA | NA |
| FKBP5 | 2 | 8 | 26 | 1 | 0,05 | 0,24 | 0,08 | 0,63 | 0,07 | 0,15 | 0,1 | 0,68 |
| TSC22D3 | 19 | 226 | 15 | 96 | 0,19 | 0,24 | 0,3 | 0,27 | 0,36 | 0,34 | 0,15 | 0,15 |

### energy metabolism

| | MoH | MoS | NeH | NeS | MoH | MoS | NeH | NeS | LyH | LyS | MyH | MyS |
|---|---|---|---|---|---|---|---|---|---|---|---|---|
| ECHDC3 | 12 | 52 | 9 | 4 | 0,16 | 0,3 | 0,1 | 0,44 | 0,07 | 0,1 | 0,22 | 0,61 |
| LPL | 1478 | 76 | 5766 | 5784 | 0,42 | 0,4 | 0,09 | 0,09 | 0,22 | 0,22 | 0,29 | 0,28 |
| SLC1A3 | 191 | 57 | 9864 | 3396 | 0,12 | 0,37 | 0,02 | 0,49 | 0,07 | 0,09 | 0,12 | 0,72 |
| BCAT1 | 4645 | 21 | 2371 | 169 | 0,2 | 0,52 | 0,04 | 0,24 | 0,06 | 0,09 | 0,2 | 0,64 |
| PFKFB2 | 10699 | 12035 | 39 | 15 | 0,06 | 0,19 | 0,07 | 0,69 | 0,09 | 0,22 | 0,09 | 0,6 |
| SIRT5 | 1711 | 86 | 3936 | 11 | 0,24 | 0,3 | 0,09 | 0,37 | 0,33 | 0,38 | 0,1 | 0,2 |

### secretory pathway

| | MoH | MoS | NeH | NeS | MoH | MoS | NeH | NeS | LyH | LyS | MyH | MyS |
|---|---|---|---|---|---|---|---|---|---|---|---|---|
| ERN1 | 1264 | 7228 | 28 | 327 | 0,16 | 0,15 | 0,39 | 0,3 | 0,34 | 0,35 | 0,17 | 0,14 |
| ERLIN1 | 41 | 40 | 4 | 67 | 0,13 | 0,35 | 0,15 | 0,37 | 0,09 | 0,14 | 0,22 | 0,55 |
| TEX2 | 24 | 19 | 3704 | 407 | 0,31 | 0,53 | 0,08 | 0,08 | 0,29 | 0,3 | 0,16 | 0,25 |
| ERGIC1 | 81 | 144 | 456 | 180 | 0,12 | 0,13 | 0,45 | 0,3 | 0,15 | 0,17 | 0,39 | 0,29 |
| GALNT2 | 831 | 381 | 3628 | 619 | 0,24 | 0,33 | 0,05 | 0,38 | 0,14 | 0,18 | 0,2 | 0,49 |
| MAN2A2 | 90 | 1257 | 107 | 122 | 0,25 | 0,19 | 0,32 | 0,25 | 0,24 | 0,23 | 0,3 | 0,23 |
| CSGALNACT2 | 848 | 101 | 1006 | 49 | 0,23 | 0,27 | 0,12 | 0,38 | 0,15 | 0,15 | 0,24 | 0,46 |
| ST6GALNAC3 | 5838 | 34 | 8908 | 34 | 0,13 | 0,23 | 0,08 | 0,56 | 0,12 | 0,17 | 0,15 | 0,55 |
| PXYLP1 | 1663 | 184 | 557 | 224 | 0,23 | 0,21 | 0,18 | 0,37 | 0,32 | 0,29 | 0,16 | 0,23 |
| B4GALT4 | 3131 | 180 | 4098 | 23 | 0,26 | 0,34 | 0,11 | 0,29 | 0,37 | 0,37 | 0,1 | 0,16 |
| TPST1 | 13 | 2 | 14 | 26 | 0,05 | 0,24 | 0,34 | 0,37 | 0,16 | 0,12 | 0,28 | 0,44 |

### sphingolipid synthesis

| | MoH | MoS | NeH | NeS | MoH | MoS | NeH | NeS | LyH | LyS | MyH | MyS |
|---|---|---|---|---|---|---|---|---|---|---|---|---|
| SPTLC2 | 822 | 83 | 6845 | 43 | 0,24 | 0,36 | 0,1 | 0,3 | 0,11 | 0,13 | 0,26 | 0,51 |
| SGMS2 | 62 | 167 | 5881 | 288 | 0,36 | 0,43 | 0,02 | 0,19 | 0,02 | 0,04 | 0,36 | 0,58 |

### extracellular matrix

| | MoH | MoS | NeH | NeS | MoH | MoS | NeH | NeS | LyH | LyS | MyH | MyS |
|---|---|---|---|---|---|---|---|---|---|---|---|---|
| ADAMTS2 | 176 | 9 | 3585 | 627 | 0,02 | 0,97 | 0 | 0 | 0,02 | 0,02 | 0,02 | 0,94 |
| THBS1 | 676 | 399 | 11753 | 322 | 0,22 | 0,77 | 0 | 0,01 | 0,01 | 0,02 | 0,22 | 0,75 |
| TIMP4 | 9121 | 64 | 2812 | 3993 | 0,21 | 0,47 | 0,14 | 0,18 | 0,27 | 0,27 | 0,16 | 0,3 |

### GPCR

| | MoH | MoS | NeH | NeS | MoH | MoS | NeH | NeS | LyH | LyS | MyH | MyS |
|---|---|---|---|---|---|---|---|---|---|---|---|---|
| GPER1 | 9 | 23 | 4898 | 3761 | 0,11 | 0,86 | 0,01 | 0,01 | 0,07 | 0,08 | 0,11 | 0,74 |
| ADORA3 | 21 | 3 | 31 | 93 | NA | NA | NA | NA | NA | NA | NA | NA |
| CXCR4 | 51 | 1451 | 3143 | 12969 | 0,23 | 0,1 | 0,56 | 0,11 | 0,44 | 0,41 | 0,12 | 0,03 |
| LTB4R | 369 | 88 | 192 | 200 | 0,18 | 0,24 | 0,17 | 0,4 | 0,15 | 0,2 | 0,23 | 0,43 |
| FPR1 | 94 | 216 | 136 | 866 | NA | NA | NA | NA | NA | NA | NA | NA |

### phagocytosis and cell motility

| | MoH | MoS | NeH | NeS | MoH | MoS | NeH | NeS | LyH | LyS | MyH | MyS |
|---|---|---|---|---|---|---|---|---|---|---|---|---|
| AMPH | 945 | 6 | 1560 | 405 | 0,1 | 0,63 | 0,03 | 0,25 | 0,15 | 0,16 | 0,09 | 0,6 |
| SRGAP1 | 1930 | 14 | 3047 | 953 | 0,29 | 0,47 | 0,11 | 0,13 | 0,23 | 0,23 | 0,22 | 0,32 |
| ARHGAP24 | 1828 | 48 | 5031 | 131 | 0,26 | 0,3 | 0,07 | 0,38 | 0,27 | 0,16 | 0,18 | 0,39 |
| RALGAPA2 | 1073 | 126 | 155 | 181 | 0,11 | 0,17 | 0,26 | 0,46 | 0,15 | 0,17 | 0,25 | 0,43 |
| SIPA1L2 | 2977 | 511 | 135 | 85 | 0,06 | 0,11 | 0,31 | 0,51 | 0,03 | 0,03 | 0,35 | 0,59 |
| RASGEF1A | 1146 | 794 | 364 | 28 | 0,1 | 0,18 | 0,3 | 0,41 | 0,28 | 0,37 | 0,14 | 0,21 |
| DAAM2 | 11255 | 47 | 21 | 2 | 0,01 | 0,21 | 0,11 | 0,68 | 0,02 | 0,06 | 0,1 | 0,82 |
| FMN1 | 823 | 5 | 4233 | 183 | 0,23 | 0,63 | 0,02 | 0,12 | 0,19 | 0,18 | 0,15 | 0,47 |
| ITSN1 | 95 | 123 | 1317 | 1993 | 0,39 | 0,53 | 0,03 | 0,05 | 0,1 | 0,1 | 0,34 | 0,47 |

### phagocytosis of apoptotic cells

| | MoH | MoS | NeH | NeS | MoH | MoS | NeH | NeS | LyH | LyS | MyH | MyS |
|---|---|---|---|---|---|---|---|---|---|---|---|---|
| MFGE8 | 5955 | 16 | 12320 | 3323 | 0,16 | 0,75 | 0,04 | 0,05 | 0,48 | 0,43 | 0,02 | 0,07 |
| VSIG4 | 1149 | 27 | 10963 | 593 | 0,02 | 0,97 | 0 | 0 | 0,01 | 0,01 | 0,03 | 0,96 |
| C1QA | 12870 | 416 | 14297 | 10001 | 0,04 | 0,95 | 0 | 0 | 0,01 | 0,01 | 0,05 | 0,93 |
| C1QB | 10207 | 359 | 10418 | 6938 | 0,02 | 0,98 | 0 | 0 | 0,01 | 0,01 | 0,02 | 0,95 |
| C1QC | 8868 | 372 | 13115 | 13272 | 0,02 | 0,98 | 0 | 0 | 0,02 | 0,02 | 0,02 | 0,95 |

RANK CORRELATION GC | EXPRESSION RATIO

### PRR / phagocytosis receptors

| | MoH | MoS | NeH | NeS | MoH | MoS | NeH | NeS | LyH | LyS | MyH | MyS |
|---|---|---|---|---|---|---|---|---|---|---|---|---|
| CLEC4E | 72 | 286 | 39 | 33 | 0,13 | 0,14 | 0,37 | 0,36 | 0 | 0,02 | 0,49 | 0,49 |
| CEACAM4 | 174 | 66 | 257 | 39 | 0,06 | 0,2 | 0,27 | 0,47 | 0 | 0,03 | 0,31 | 0,65 |

### endocytosis receptor

| | MoH | MoS | NeH | NeS | MoH | MoS | NeH | NeS | LyH | LyS | MyH | MyS |
|---|---|---|---|---|---|---|---|---|---|---|---|---|
| CD163 | 16 | 7 | 2093 | 51 | 0,14 | 0,56 | 0 | 0,29 | 0 | 0,01 | 0,14 | 0,85 |

### IL1 and IL18 receptors and signaling

| | MoH | MoS | NeH | NeS | MoH | MoS | NeH | NeS | LyH | LyS | MyH | MyS |
|---|---|---|---|---|---|---|---|---|---|---|---|---|
| IRAK3 | 25 | 46 | 73 | 16 | 0,19 | 0,25 | 0,09 | 0,48 | 0,01 | 0,04 | 0,26 | 0,69 |
| IL1R2 | 8 | 4 | 3 | 5 | 0 | 0,1 | 0,11 | 0,79 | 0 | 0,04 | 0,11 | 0,85 |
| IL1R1 | 2191 | 2753 | 1140 | 54 | 0,03 | 0,08 | 0,3 | 0,58 | 0,03 | 0,04 | 0,31 | 0,62 |
| IL18R1 | 4923 | 367 | 1843 | 6 | 0 | 0,07 | 0,13 | 0,8 | 0,12 | 0,19 | 0,09 | 0,6 |
| IL18RAP | 3439 | 3679 | 6244 | 37 | 0 | 0,05 | 0,24 | 0,71 | 0,16 | 0,13 | 0,17 | 0,53 |

### receptors JAK/STAT

| | MoH | MoS | NeH | NeS | MoH | MoS | NeH | NeS | LyH | LyS | MyH | MyS |
|---|---|---|---|---|---|---|---|---|---|---|---|---|
| IFNGR1 | 1758 | 60 | 137 | 81 | 0,16 | 0,2 | 0,31 | 0,33 | 0,08 | 0,08 | 0,39 | 0,45 |
| IL13RA1 | 34 | 330 | 1715 | 162 | 0,2 | 0,15 | 0,42 | 0,23 | 0,03 | 0,04 | 0,58 | 0,36 |

### receptor kinases

| | MoH | MoS | NeH | NeS | MoH | MoS | NeH | NeS | LyH | LyS | MyH | MyS |
|---|---|---|---|---|---|---|---|---|---|---|---|---|
| FLT3 | 7 | 1 | 12228 | 87 | 0,27 | 0,69 | 0,01 | 0,03 | 0,04 | 0,07 | 0,25 | 0,64 |
| INSR | 50 | 475 | 3880 | 280 | 0,52 | 0,43 | 0,02 | 0,03 | 0,06 | 0,11 | 0,45 | 0,38 |
| ACVR1B | 137 | 129 | 128 | 100 | 0,22 | 0,3 | 0,08 | 0,4 | 0,15 | 0,15 | 0,21 | 0,49 |

### intracellular signaling, misc.

| | MoH | MoS | NeH | NeS | MoH | MoS | NeH | NeS | LyH | LyS | MyH | MyS |
|---|---|---|---|---|---|---|---|---|---|---|---|---|
| IRS2 | 5 | 274 | 2 | 179 | 0,22 | 0,31 | 0,26 | 0,21 | 0,23 | 0,16 | 0,3 | 0,32 |
| GRB10 | 7670 | 10 | 169 | 3 | 0,04 | 0,21 | 0,06 | 0,69 | 0,04 | 0,06 | 0,08 | 0,81 |
| PTEN | 57 | 37 | 426 | 308 | 0,1 | 0,15 | 0,38 | 0,36 | 0,1 | 0,12 | 0,38 | 0,4 |
| ITPKC | 39 | 35 | 4785 | 242 | 0,12 | 0,27 | 0,16 | 0,46 | 0,17 | 0,19 | 0,18 | 0,46 |
| DDIT4 | 193 | 22 | 734 | 9951 | 0,12 | 0,14 | 0,46 | 0,29 | 0,34 | 0,37 | 0,17 | 0,12 |
| SESN1 | 2690 | 25 | 5779 | 1126 | 0,28 | 0,46 | 0,12 | 0,14 | 0,36 | 0,49 | 0,06 | 0,09 |
| PICK1 | 4 | 33 | 5750 | 4082 | 0,31 | 0,47 | 0,12 | 0,1 | 0,31 | 0,35 | 0,15 | 0,19 |
| VSIR | 209 | 2623 | 8 | 40 | 0,19 | 0,19 | 0,31 | 0,31 | 0,04 | 0,06 | 0,45 | 0,45 |

### SPMs, prostaglandins, leukotrienes

| | MoH | MoS | NeH | NeS | MoH | MoS | NeH | NeS | LyH | LyS | MyH | MyS |
|---|---|---|---|---|---|---|---|---|---|---|---|---|
| ALOX15B | 15 | 12 | 3025 | 230 | 0,35 | 0,6 | 0,03 | 0,03 | 0,1 | 0,1 | 0,3 | 0,5 |
| HPGD | 161 | 24 | 14555 | 137 | 0,01 | 0,07 | 0,01 | 0,91 | 0,06 | 0,09 | 0,01 | 0,84 |
| ALOX5AP | 1172 | 140 | 2409 | 419 | 0,05 | 0,12 | 0,43 | 0,41 | 0,07 | 0,11 | 0,39 | 0,43 |

### angiogenesis and more

| | MoH | MoS | NeH | NeS | MoH | MoS | NeH | NeS | LyH | LyS | MyH | MyS |
|---|---|---|---|---|---|---|---|---|---|---|---|---|
| RNASE1 | 9996 | 238 | 4433 | 490 | 0,1 | 0,85 | 0,03 | 0,03 | 0,09 | 0,09 | 0,1 | 0,72 |
| ANG | 1368 | 112 | 7807 | 354 | 0,37 | 0,56 | 0,04 | 0,04 | 0,13 | 0,13 | 0,3 | 0,44 |
| RNASE4 | 183 | 158 | 7936 | 222 | 0,32 | 0,33 | 0,17 | 0,17 | 0,3 | 0,3 | 0,2 | 0,2 |

### miscellaneous

| | MoH | MoS | NeH | NeS | MoH | MoS | NeH | NeS | LyH | LyS | MyH | MyS |
|---|---|---|---|---|---|---|---|---|---|---|---|---|
| MACIR | 10195 | 31 | 176 | 18 | 0,11 | 0,25 | 0,13 | 0,51 | 0,22 | 0,3 | 0,11 | 0,37 |
| STS | 1231 | 69 | 613 | 90 | 0,37 | 0,37 | 0,11 | 0,15 | 0,15 | 0,17 | 0,32 | 0,35 |
| MAOA | 4478 | 41 | 2403 | 146 | 0,16 | 0,51 | 0,09 | 0,25 | 0,2 | 0,2 | 0,14 | 0,45 |
| PRL | 9253 | 49 | 11369 | 1938 | 0,22 | 0,35 | 0,14 | 0,29 | 0,27 | 0,28 | 0,16 | 0,29 |
| APH1B | 8592 | 317 | 330 | 30 | 0,21 | 0,27 | 0,23 | 0,28 | 0,22 | 0,24 | 0,24 | 0,3 |
| OLAH | 4750 | 42 | 2272 | 14 | 0,01 | 0,04 | 0,01 | 0,94 | 0,02 | 0,04 | 0,02 | 0,92 |
| ARG1 | 12397 | 2190 | 266 | 21 | 0,01 | 0,08 | 0,06 | 0,85 | 0,02 | 0,09 | 0,06 | 0,84 |
