## Supplementary material for "Glucocorticoid-driven gene expression in circulating monocytes and neutrophils in health and severe inflammation": Figure S11

# A

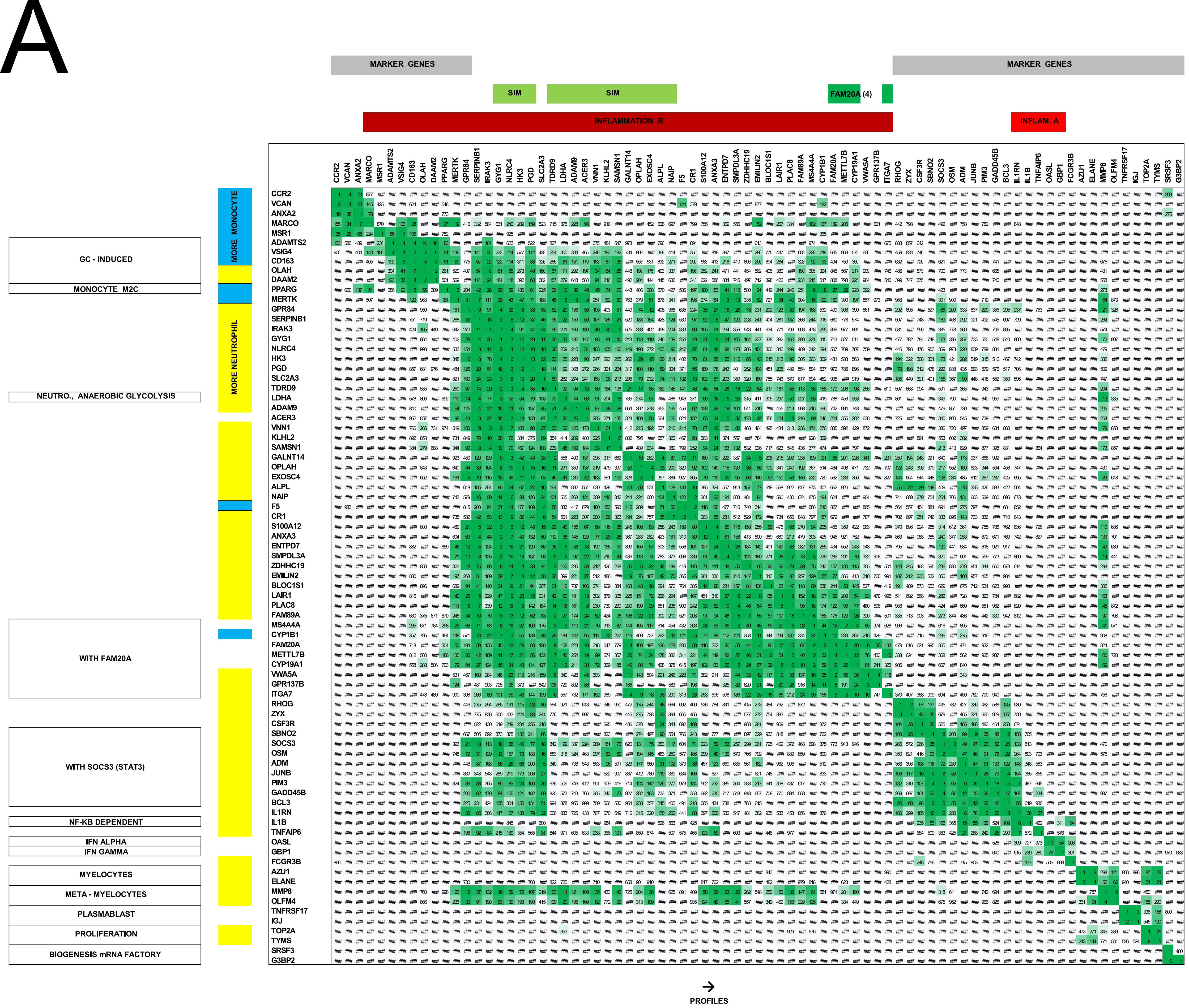


**SET SIM : GYG1, NLRC4, HK3, PGD, TDRD9, LDHA, ADAM9, ACER3, VNN1, KLHL2, SAMSN1, GALNT14, OPLAH, EXOSC4, ALPL, NAIP**

**SET FAM20A (4) : FAM20A, METTL7B, ITGA7, CYP19A1**

# B

| | ID_REF | Log2 FC mild 1 | Log2 FC sev 1 | Log10 pval mild 1 | Log10 pval sev 1 | | Log2 FC sev 2 | Log10 pval sev 2 |
|---|---|---|---|---|---|---|---|---|
| LDHA | ILMN_1807106 | 0,19 | 1,05 | -13,1 | -14,7 | LDHA | 1,46 | -13,1 |
| ADAM9 | ILMN_1727524 | 0,02 | 0,81 | -11,5 | -8,43 | ADAM9 | 1,25 | -16,4 |
| VNN1 | ILMN_1674574 | 0,03 | 2,8 | -1,49 | -36,4 | VNN1 | 2,65 | -12,9 |
| KLHL2 | ILMN_1701837 | 0,05 | 1,73 | -2,09 | -14,9 | KLHL2 | 1,21 | -14,4 |
| SAMSN1 | ILMN_2171289 | 0,03 | 1,32 | -6,97 | -16,4 | SAMSN1 | 2,06 | -6,04 |
| NAIP | ILMN_2260082 | 0,2 | 1,01 | -25,2 | -6,98 | NAIP | 1,49 | -15,8 |
| ALPL | ILMN_1701603 | 0,21 | NA | -1,42 | NA | ALPL | 1,76 | -19 |
| GALNT14 | ILMN_1722524 | 0,01 | 1,61 | -2,32 | -11,5 | GALNT14 | 2,27 | -16,1 |
| OPLAH | ILMN_1711030 | 0,06 | 2,94 | -4,84 | -33,2 | OPLAH | 2,05 | -16 |
| EXOSC4 | ILMN_1745271 | 0,03 | 1,8 | -6,14 | -28,6 | EXOSC4 | 1,67 | -11,3 |
| GYG1 | ILMN_2230862 | 0,55 | 2,63 | -19,9 | -27,5 | GYG1 | 2,28 | -14,5 |
| NLRC4 | ILMN_1796976 | 0,17 | NA | -29,3 | NA | NLRC4 | 1,71 | -21,2 |
| HK3 | ILMN_1670302 | 0,48 | 2,26 | -33,9 | -31 | HK3 | 1,84 | -18,2 |
| PGD | ILMN_1794165 | 0,19 | 1,79 | -5,8 | -27,2 | PGD | 1,49 | -18,6 |
| ACER3 | ILMN_3237774 | 0,03 | NA | -11,1 | NA | ACER3 | 1,2 | -12,5 |
| TDRD9 | ILMN_2169490 | 0,04 | 3,34 | -7,63 | -40,5 | TDRD9 | 2,35 | -10,3 |
| VWA5A | ILMN_1764769 | 0,02 | NA | -5,18 | NA | VWA5A | 1,05 | -6,59 |
| GPR137B | ILMN_2121816 | 0,06 | 0,7 | -20,6 | -5,85 | GPR137B | 0,69 | -3,25 |
| ITGA7 | ILMN_1791409 | 0 | 0,58 | -0,15 | -11,3 | ITGA7 | 3,08 | -7,67 |
| CYP19A1 | ILMN_2387860 | 0 | 1,37 | -0,84 | -8,91 | CYP19A1 | 4,8 | -3,2 |
| FAM20A | ILMN_1812091 | 0,02 | 1,15 | -7,3 | -15,1 | FAM20A | 3,26 | -13,9 |
| METTL7B | ILMN_2064725 | 0,02 | 2,52 | -8,32 | -17,5 | METTL7B | 3,57 | -8,45 |

# C

| | Mo control | Ne control | Mo IFN | Ne IFN | Mo sepsis | Ne sepsis |
|---|---|---|---|---|---|---|
| LDHA | 0,16 | 0,05 | 0,17 | 0,03 | 0,31 | 0,27 |
| ADAM9 | 0,19 | 0,04 | 0,15 | 0,04 | 0,39 | 0,19 |
| VNN1 | 0,07 | 0,13 | 0,03 | 0,05 | 0,16 | 0,56 |
| KLHL2 | 0,06 | 0,18 | 0,05 | 0,11 | 0,12 | 0,47 |
| SAMSN1 | 0,11 | 0,08 | 0,12 | 0,09 | 0,2 | 0,4 |
| NAIP | 0,08 | 0,07 | 0,07 | 0,08 | 0,19 | 0,51 |
| ALPL | 0 | 0,19 | 0 | 0,11 | 0,04 | 0,65 |
| GALNT14 | 0,01 | 0,12 | 0,01 | 0,05 | 0,07 | 0,75 |
| OPLAH | 0,06 | 0,1 | 0,04 | 0,04 | 0,13 | 0,62 |
| EXOSC4 | 0,11 | 0,08 | 0,09 | 0,07 | 0,16 | 0,48 |
| GYG1 | 0,06 | 0,05 | 0,06 | 0,05 | 0,18 | 0,6 |
| NLRC4 | 0,05 | 0,08 | 0,06 | 0,06 | 0,13 | 0,63 |
| HK3 | 0,14 | 0,05 | 0,12 | 0,07 | 0,21 | 0,41 |
| PGD | 0,11 | 0,11 | 0,09 | 0,08 | 0,22 | 0,39 |
| ACER3 | 0,19 | 0,02 | 0,21 | 0,01 | 0,29 | 0,29 |
| TDRD9 | 0,09 | 0,02 | 0,05 | 0,02 | 0,25 | 0,57 |
| VWA5A | 0,08 | 0,01 | 0,08 | 0,01 | 0,25 | 0,57 |
| GPR137B | 0,15 | 0,01 | 0,16 | 0,01 | 0,34 | 0,33 |
| ITGA7 | 0,04 | 0,02 | 0,05 | 0,01 | 0,25 | 0,64 |
| CYP19A1 | 0,01 | 0,01 | 0,01 | 0,01 | 0,29 | 0,67 |
| FAM20A | 0,01 | 0,01 | 0,01 | 0,01 | 0,56 | 0,38 |
| METTL7B | 0,04 | 0,02 | 0,06 | 0,02 | 0,64 | 0,21 |

# D

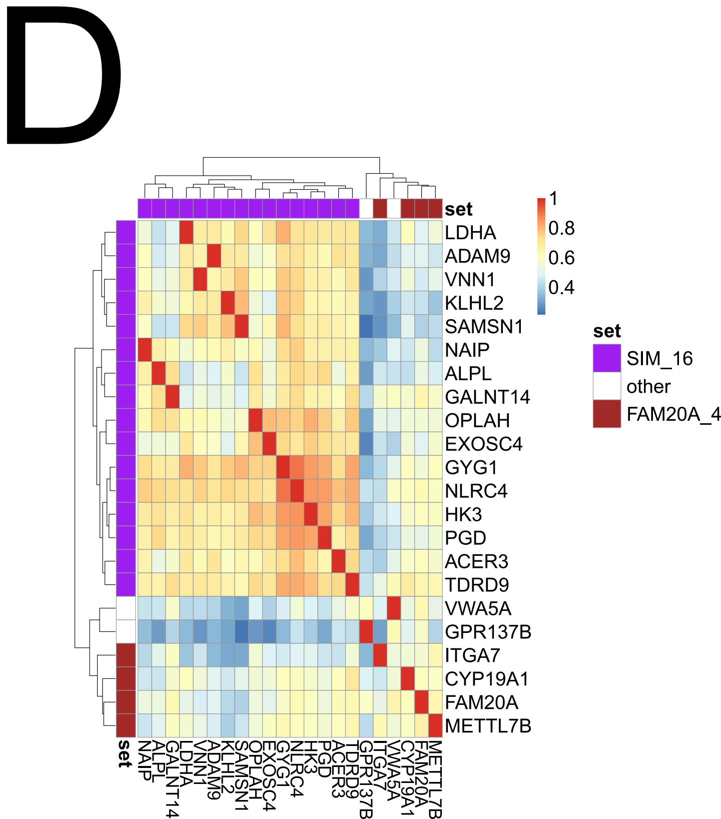
